# Completeness, consistency and analytical usability of routine data for preterm and small newborn care in Western Kenya: a multicentre mixed-methods study

**DOI:** 10.64898/2026.11.16.26368243

**Authors:** Morris Senghor Shisanya, Lucy Natecho Namusonge, Catherine Mutonyi Simiyu, Tecla Cecilia Kirwa, Everlyne Nyanchera Morema

**Author notes:** Corresponding author: Morris Senghor Shisanya, Department of Community Health and Mental Health Nursing, School of Nursing, Kibabii University, Bungoma, Kenya, (alternative).

## Abstract

**Objectives:** To assess the completeness, consistency, plausibility, uniqueness and analytical usability of routine maternity and newborn records used for preterm and small newborn care, and how facility health-information-system readiness and working conditions explain these patterns.

**Design:** Multicentre cross-sectional mixed-methods study of routine health data with embedded explanatory qualitative interviews.

**Setting Twelve health facilities:** (levels 3–5) in Bungoma and one neighbouring county, Western Kenya.

**Participants 349:** maternity-register and 291 newborn records for deliveries from July 2024 to May 2026; 10 facility readiness assessments; two provider key informant interviews.

**Primary and secondary outcome measures:** Variable-and record-level completeness from original uncleaned abstractions; prespecified consistency and plausibility rule violations; duplicates; agreement between recorded and rule-derived preterm, low-birth-weight (LBW) and very-low-birth-weight classifications.

**Results:** Median record completeness was 100% (IQR 94.7–100) for register and 93.5% (IQR 84.8–100) for newborn records; a complete core record was present in 175/349 (50.1%, 95% CI 44.9 to 55.4) and 77/291 (26.5%, 95% CI 21.7 to 31.8) respectively. No maternal age, gestational age, birth weight or Apgar value was implausible, but chronological errors were common: recorded and date-derived length of stay differed by >1 day in 92/225 (40.9%) and admission preceded birth in 35/257 (13.6%). Agreement with rule-derived classification was high for very low birth weight (kappa 0.89, 95% CI 0.83 to 0.95) but moderate for LBW (kappa 0.53, 95% CI 0.32 to 0.73; McNemar p=0.001). Complete documentation was more likely with three or more record sources (adjusted prevalence ratio 1.80, 95% CI 1.05 to 3.08) and less likely for newborns who died (0.46, 95% CI 0.25 to 0.84). Providers described documentation competing with workload.

**Conclusions:** Identification, gestational age, birth weight and disposition were documented sufficiently for classification and mortality analysis; timing fields, maternal history and transport variables were not. Preterm-birth prevalence was not estimable without an all-delivery denominator.

**Strengths and limitations of this study:**

- All completeness, consistency and plausibility estimates were computed from the original, unedited abstractions, without imputation or correction, so that the records were assessed as they exist in routine practice.
- Three routine data sources from twelve facilities were assessed against prespecified rules, and every dataset was verified cell by cell against the original electronic export before analysis.
- Recalculated classifications served only as a rules-based consistency reference; no independent clinical gold standard (such as direct observation or re-measurement) was available, so accuracy could not be assessed.
- Only ten facilities contributed readiness data, so facility-level correlations are exploratory and imprecise, and no complex facility-level model was fitted.
- The explanatory qualitative component comprised two provider interviews at the county referral hospital and therefore illustrates rather than represents documentation practice across all facility levels.

## Introduction

Routine health information is the operational backbone of newborn care. Facility registers, admission records, monitoring charts and discharge summaries determine what clinicians know at the bedside, what audit committees can review, and what counties can count. The World Health Organization Standards for improving the quality of care for small and sick newborns make this explicit: actionable health information systems are one of eight standards, on the reasoning that care cannot be improved if it cannot be described.[1] The Every Newborn Action Plan measurement roadmap reaches the same conclusion from the measurement side, identifying care of small and sick newborns as the area where routine data are weakest.[2]

An estimated 13.4 million babies were born preterm in 2020, with sub-Saharan Africa and southern Asia carrying most of the burden and most of the associated mortality,[3,4] and Kenya’s neonatal mortality rate stands at 21 deaths per 1,000 live births.[5] Preterm and small newborns place unusually heavy demands on documentation. Their classification depends on two measurements — gestational age and birth weight — that are themselves difficult to obtain reliably where last menstrual period rather than early ultrasound remains the dominant dating method.[6,7,8] Their management is time-critical, so the value of a record depends on timestamps as much as on values. Their outcomes are frequently determined within hours, so documentation opportunities are unevenly distributed between newborns who die early and those who survive to discharge. Each feature creates a distinct way for a record to fail.

Where routine records are adequate they support surveillance, clinical audit, perinatal death review, quality improvement and planning without the cost of bespoke data collection.[9] Where they are not, the consequences propagate silently: indicators are computed from unstable denominators, mortality audits proceed without the fields needed to identify modifiable factors, and research inherits errors invisible at analysis. A systematic review of routine facility data for newborn indicators in low-and middle-income countries found completeness ranging from 0% to 100% across registers and case notes, with only four studies reporting internal consistency, and concluded that the evidence base is thin relative to the investment made in these systems;[10] a recent four-country assessment reached similar conclusions.[11]

Kenya has invested substantially in routine health information. A nationwide evaluation of the Kenya Health Information System (KHIS), built on DHIS2, reported a mean facility-quarter completeness score of 61.1% alongside a consistency score of 80.0%,[12] and qualitative work across 15 counties described limited analytical capacity as a persistent constraint on data use,[13] within a wider Eastern and Southern African pattern of uneven performance.[14] Newborn care has benefited from the Clinical Information Network (CIN), a KEMRI-coordinated learning health system in which structured newborn admission records feed a common platform supporting audit and feedback.[15,16,17] Yet capturing neonatal data for global monitoring remains difficult even where systems exist,[18] hospital information systems often cannot support evidence-informed decisions,[19] and verification of childbirth registers in Western Kenya has documented substantial gaps.[20]

What remains largely unexamined is the quality of the specific records a county would use to assess its own preterm and small newborn care. Most assessments concentrate on registers or aggregate DHIS2 reports; few examine individual newborn case notes, and fewer still assess linked maternity and newborn records together with the facility information-system capacity that produces them.[10] This matters because the fields that fail are not the fields most often audited: completeness of birth weight says little about whether admission time was recorded, whether the two places a birth weight is written agree, or whether length of stay can be computed.

We therefore assessed the completeness, internal consistency, plausibility, uniqueness and analytical usability of routine maternity and newborn records across twelve facilities in Western Kenya, and examined how facility information-system readiness and frontline working conditions help explain these patterns. Objectives were to quantify variable-level and record-level completeness; assess consistency among logically related variables; assess agreement between recorded and rule-derived classifications; quantify implausible values, chronological errors and duplicates; determine which indicators the data support; examine facility variation and its relationship with readiness; identify record-level predictors of complete documentation; and integrate provider accounts with the quantitative findings.

## Methods

### Study design and reporting

This was a multicentre cross-sectional assessment of routinely collected health data with an embedded explanatory qualitative component, nested within a mixed-methods study of health-system readiness and quality of care for preterm births in Western Kenya. Reporting follows the RECORD extension to STROBE,[21,22] with selected COREQ principles applied to the embedded provider component (completed RECORD checklist, online supplemental file 1).[23]

### Setting

Bungoma County operates a tiered public health system comprising health centres (level 3), sub-county hospitals (level 4) and a county referral hospital (level 5). Twelve public facilities contributed data: eleven in Bungoma County and the county referral hospital of neighbouring Busia County. Ten underwent a facility readiness assessment, nine (all in Bungoma) contributed patient-level records, and seven contributed both. All provided maternity services; of the ten assessed facilities, seven had a newborn unit and two a neonatal intensive care unit. The Bungoma county referral hospital participates in the Clinical Information Network, uses REDCap-based abstraction from structured paper forms and receives partner-supported dashboard reporting.

### Data sources

Three structured instruments, approved before data collection, were administered by trained research assistants using Kobo Toolbox on tablets. Tool 1 abstracted maternity-register entries for recorded deliveries (57 variables). Tool 2 assessed facility readiness across 11 domains on a three-point scale (0 = not available; 1 = available but non-functional or inconsistently used; 2 = fully available and functional), with free-text fields on strengths, gaps and recommendations (122 variables). Tool 3 abstracted individual newborn clinical records, including a 31-item patient-level quality score (PLQS) covering eight care domains scored on the same scale with an explicit ‘not applicable’ option (186 variables). Data were collected between 4 May and 26 June 2026. Two provider key informant interviews were conducted at the county referral hospital, with the consultant paediatrician and head of the newborn unit and with the newborn-unit nurse manager; both were audio-recorded with consent and transcribed verbatim.

### Data management

Because this study assesses the quality of the original documentation, all completeness, consistency and plausibility estimates were computed from the abstractions as entered, without imputation, correction or recoding of missing values.

Cleaned datasets were used only for facility-name standardisation, duplicate identification and unique record counts. Before analysis, each dataset was verified cell by cell against the original Kobo Toolbox export, and all analyses were re-run to confirm that outputs were reproducible.

### Facility standardisation and duplicate resolution

Facility names were free-text and appeared in numerous forms; a master crosswalk mapping every observed alias to a standard name, KHIS code, sub-county and level was applied to both patient-level datasets. Potential duplicates were identified using facility, record identifier, date and newborn characteristics, and classified as confirmed duplicate, true multiple birth or distinct record. Within confirmed duplicate groups the record with the highest weighted completeness was retained and singular non-conflicting values recovered from complementary records; contradictory values were never silently merged and remain visible in the duplicate-resolution log. Documented multiple births were retained as separate newborns.

### Data-quality definitions

Five dimensions were assessed. Completeness was the presence of usable information in a required field; a value counted as documented only if a usable entry was present, with blanks and explicit ‘not recorded’, ‘unknown’ or ‘not documented’ entries both counted as not documented but tabulated separately, since they carry different implications for instrument design. Internal consistency was agreement between logically related variables; plausibility whether values, dates and combinations fell within defensible limits; uniqueness whether each record represented one distinct delivery or newborn; and analytical usability whether the data contained enough information to compute relevant indicators. Accuracy was not assessed, because no independent gold standard such as direct observation or re-measurement was collected.

### Completeness assessment

Nineteen core register and 34 core newborn variables were prespecified across documentation domains (identification; maternal demographics or maternal and obstetric information; antenatal and obstetric data; newborn baseline; entry and referral; admission assessment; outcome; discharge). For each, the eligible denominator, number documented, number blank, number explicitly marked not recorded, and completeness with a Wilson 95% confidence interval were computed. Structurally inapplicable items were excluded from denominators: prematurity category only where gestational age was present; referring facility level only among referred-in newborns; discharge weight and feeding at discharge only among newborns discharged alive.

Record-level completeness was the proportion of applicable core fields documented, computed overall and by domain; a complete core record required every applicable core field. Distance or travel time was retained in the variable-level table but excluded from the newborn record-level index, being documented in fewer than 1% of records.

### Consistency, plausibility and agreement analysis

Sixteen register rules and 25 newborn rules were prespecified and are listed in online supplemental table 1; construction followed the WHO data quality review framework and the PRISM approach.[24,25] Working plausibility ranges were maternal age 10–55 years, gestational age 20–44 weeks, birth weight 300–6,000 g, Apgar 0–10, gravidity 1–20, parity 0–19 and length of stay 0–180 days. Gestational age and birth weight were treated as incompatible if gestational age was below 28 weeks with weight of at least 2,000 g, or at least 34 weeks with weight below 800 g.

Recorded preterm, low-birth-weight (LBW) and very-low-birth-weight (VLBW) classifications were compared with classifications recalculated from the underlying gestational age and birth weight. The recalculated classification is a rules-based consistency reference, not an independent clinical gold standard, and results are reported as agreement rather than diagnostic validity. For each comparison we report the cross-tabulation, percentage agreement, positive and negative agreement, Cohen’s kappa with a 95% confidence interval and an exact McNemar test. Where the same quantity was captured in two fields (gestational age; birth weight) we report exact agreement, median absolute difference, the proportion within tolerance (1 week; 50 g) and Bland–Altman limits of agreement.[26] Correlation was not used as evidence of agreement.

### Analytical usability

A fitness-for-analysis matrix listed analyses commonly required for monitoring and research on preterm and small newborns, each classified as fully supported, supported with limitations, or not currently supported, with a stated reason. No numerical usability index was created.

### Facility, HIS and predictor analyses

Facility readiness scores were recalculated from observed items. Health-information-system (HIS) readiness comprised four items (newborn registers, gestational-age and birth-weight documentation, perinatal death audits, KHIS/DHIS2 reporting); quality-improvement (QI) and governance readiness comprised four items (active QI team, review meetings, use of audit data, supportive supervision). Facility-level completeness, inconsistency rate and duplicate rate were linked to these scores using the audited facility identifier. Because only ten facilities were assessed and seven contributed both readiness and patient-level data, relationships were examined using facility profiles, scatterplots and Spearman correlation with 95% confidence intervals, and are reported as exploratory and ecological. No facility-level multivariable model was fitted.

Record-level predictors of a complete core record were examined separately for each instrument using modified Poisson regression with robust variance to estimate prevalence ratios.[27] Covariates were selected conceptually rather than by stepwise procedures: the register model included facility level, period of delivery and birth outcome; the newborn model included facility level, ward or unit, number of record sources consulted, mode of entry, disposition and period of birth. A separate model restricted to facilities with a readiness assessment estimated associations with HIS and QI readiness per 25 percentage points. All completeness outcomes were derived from original documentation only.

### Missing data

Missingness is the outcome here rather than a nuisance, and was never imputed. Blank entries, explicit ‘not recorded’, ‘unknown’, ‘not assessed’ and structurally inapplicable responses were distinguished throughout. Complete-case analysis was used for all regression models, and sensitivity analyses (online supplemental table 5) include one in which any entry, including an explicit ‘not recorded’, was counted as documented.

### Qualitative analysis

Provider transcripts and Tool 2 free-text responses were analysed using the framework method.[28] A deductive codebook developed from the study instruments covered routine documentation, paper registers, electronic systems, dashboards, workload and staffing, documentation burden, discharge records, weekly and monthly audits, perinatal death review, feedback and mentorship, and use of data for quality improvement; inductive codes were added as they emerged. Two analysts coded independently and reconciled by consensus. The component is explanatory: with two interviews, both from the county referral hospital, no claim of thematic saturation or representativeness across facility levels is made. Providers are identified as Provider KII 1 (consultant paediatrician and head of the newborn unit) and Provider KII 2 (newborn-unit nurse manager). All quotations are verbatim.

### Mixed-method integration

Datasets were analysed separately and then integrated using a joint display pairing each quantitative finding with its facility information-system context, provider explanation, clinical implication and corrective action, classified as convergence, complementarity, expansion or discordance. Qualitative evidence was not forced to explain findings for which no supporting account existed.

### Statistical methods

All tests were two-sided with an alpha of 0.05 and estimates are reported with 95% confidence intervals, emphasising effect estimates over p-values. Binomial proportions use Wilson intervals;[29] kappa standard errors follow the Fleiss method;[30] McNemar tests are exact; Spearman confidence intervals use the Fisher z transformation. Analyses were conducted in Python 3 (pandas, NumPy, SciPy).

### Patient and public involvement

There was no patient or public involvement in the design, conduct, reporting or dissemination of this study.

### Ethics and research authorisations

The study was approved by the Masinde Muliro University of Science and Technology Institutional Scientific and Ethics Review Committee (MMUST-ISERC). Retrospective abstraction of routine records was approved without individual patient consent but facility in charge assent to conduct the abstraction. No direct patient identifiers were carried into the analytical datasets; facility and newborn identifiers were used only for duplicate resolution and are not reported. Providers gave informed consent to participate and to be audio-recorded. All reporting is aggregated and quotations are anonymised. Full approval details appear under Declarations.

## Results

### Study records and facilities

The three instruments yielded 349 maternity-register records, 291 newborn clinical records and 10 facility readiness assessments. Patient-level records came from nine Bungoma facilities (one level 5 county referral hospital, six level 4 hospitals and two level 3 health centres); readiness assessments covered ten facilities, including the Busia county referral hospital, and seven facilities contributed both. Recorded delivery dates spanned 15 July 2024 to 21 May 2026. Characteristics of the data sources, facilities and records are shown in table 1.

**Table 1.**
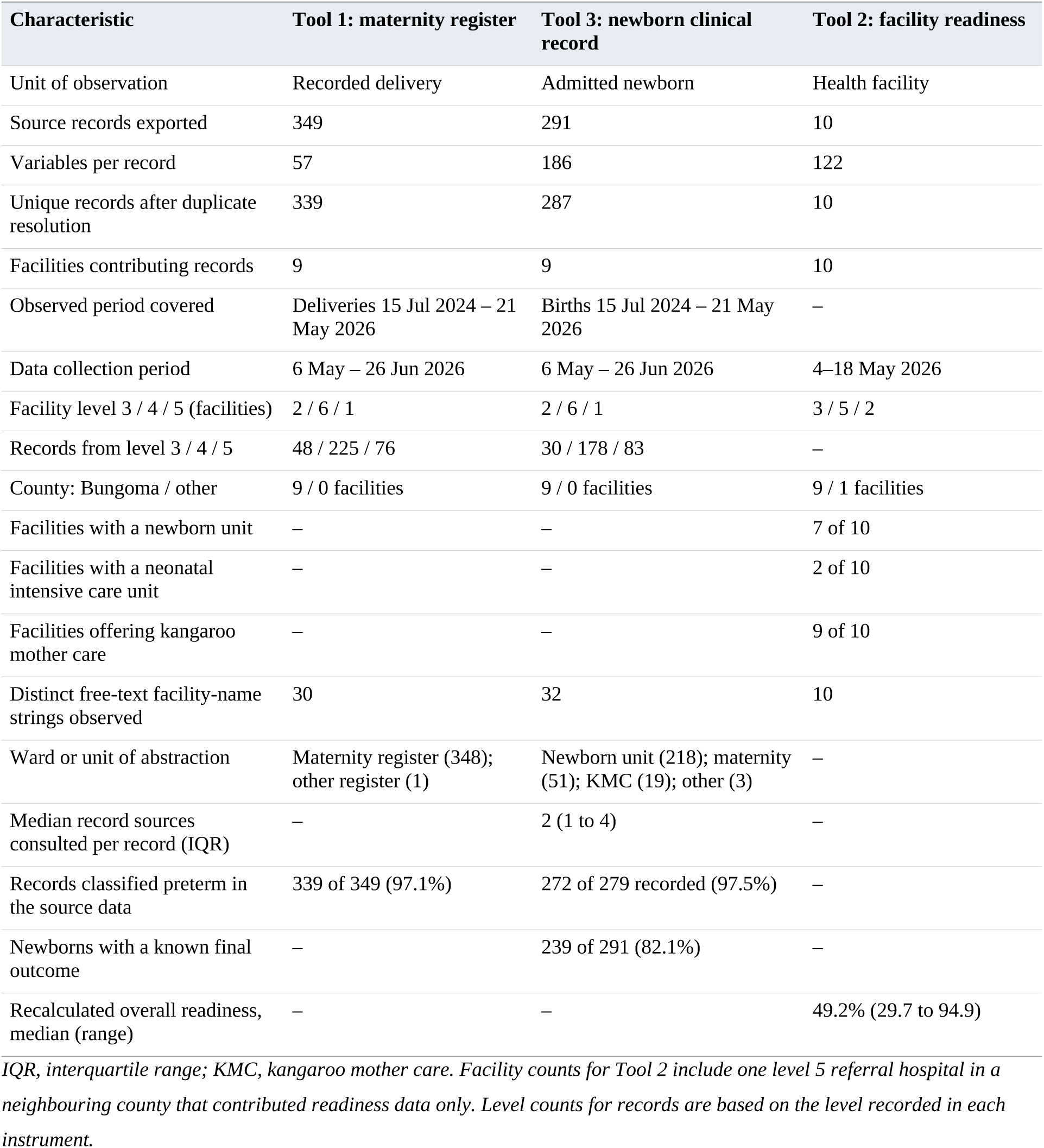
Characteristics of data sources, facilities and records.

| **Characteristic** | **Tool 1: maternity register** | **Tool 3: newborn clinical record** | **Tool 2: facility readiness** |
|---|---|---|---|
| Unit of observation | Recorded delivery | Admitted newborn | Health facility |
| Source records exported | 349 | 291 | 10 |
| Variables per record | 57 | 186 | 122 |
| Unique records after duplicate resolution | 339 | 287 | 10 |
| Facilities contributing records | 9 | 9 | 10 |
| Observed period covered | Deliveries 15 Jul 2024 – 21 May 2026 | Births 15 Jul 2024 – 21 May 2026 | – |
| Data collection period | 6 May – 26 Jun 2026 | 6 May – 26 Jun 2026 | 4–18 May 2026 |
| Facility level 3 / 4 / 5 (facilities) | 2 / 6 / 1 | 2 / 6 / 1 | 3 / 5 / 2 |
| Records from level 3 / 4 / 5 | 48 / 225 / 76 | 30 / 178 / 83 | – |
| County: Bungoma / other | 9 / 0 facilities | 9 / 0 facilities | 9 / 1 facilities |
| Facilities with a newborn unit | – | – | 7 of 10 |
| Facilities with a neonatal intensive care unit | – | – | 2 of 10 |
| Facilities offering kangaroo mother care | – | – | 9 of 10 |
| Distinct free-text facility-name strings observed | 30 | 32 | 10 |
| Ward or unit of abstraction | Maternity register (348); other register (1) | Newborn unit (218); maternity (51); KMC (19); other (3) | – |
| Median record sources consulted per record (IQR) | – | 2 (1 to 4) | – |
| Records classified preterm in the source data | 339 of 349 (97.1%) | 272 of 279 recorded (97.5%) | – |
| Newborns with a known final outcome | – | 239 of 291 (82.1%) | – |
| Recalculated overall readiness, median (range) | – | – | 49.2% (29.7 to 94.9) |

*IQR, interquartile range; KMC, kangaroo mother care. Facility counts for Tool 2 include one level 5 referral hospital in a neighbouring county that contributed readiness data only. Level counts for records are based on the level recorded in each instrument.*

### Data verification and uniqueness

Cell-by-cell verification against the original electronic exports found no discrepancy in any dataset (19,950 register, 1,342 facility-assessment and 54,312 newborn cells), and no record or variable was added or lost.

Facility identification was the single largest threat to record linkage. Thirty distinct facility-name strings in the register dataset and 32 in the newborn dataset resolved to nine facilities, and 15 sub-county spellings to seven sub-counties; one facility appeared under ten written forms. Ten duplicate groups were identified in the register dataset, of which nine were confirmed duplicates (10 records excluded) and one a documented twin pair correctly retained as two newborns; four confirmed duplicate groups were identified in the newborn dataset (4 records excluded). Three records in each dataset were reconciled by recovering singular non-conflicting values, leaving 339 unique register and 287 unique newborn records (figure 1; online supplemental table 4). Duplicates were concentrated in three facilities and reached 10.0% of register records at one health centre. All exclusions remain reversible through the record-lineage file.

**Figure 1.**
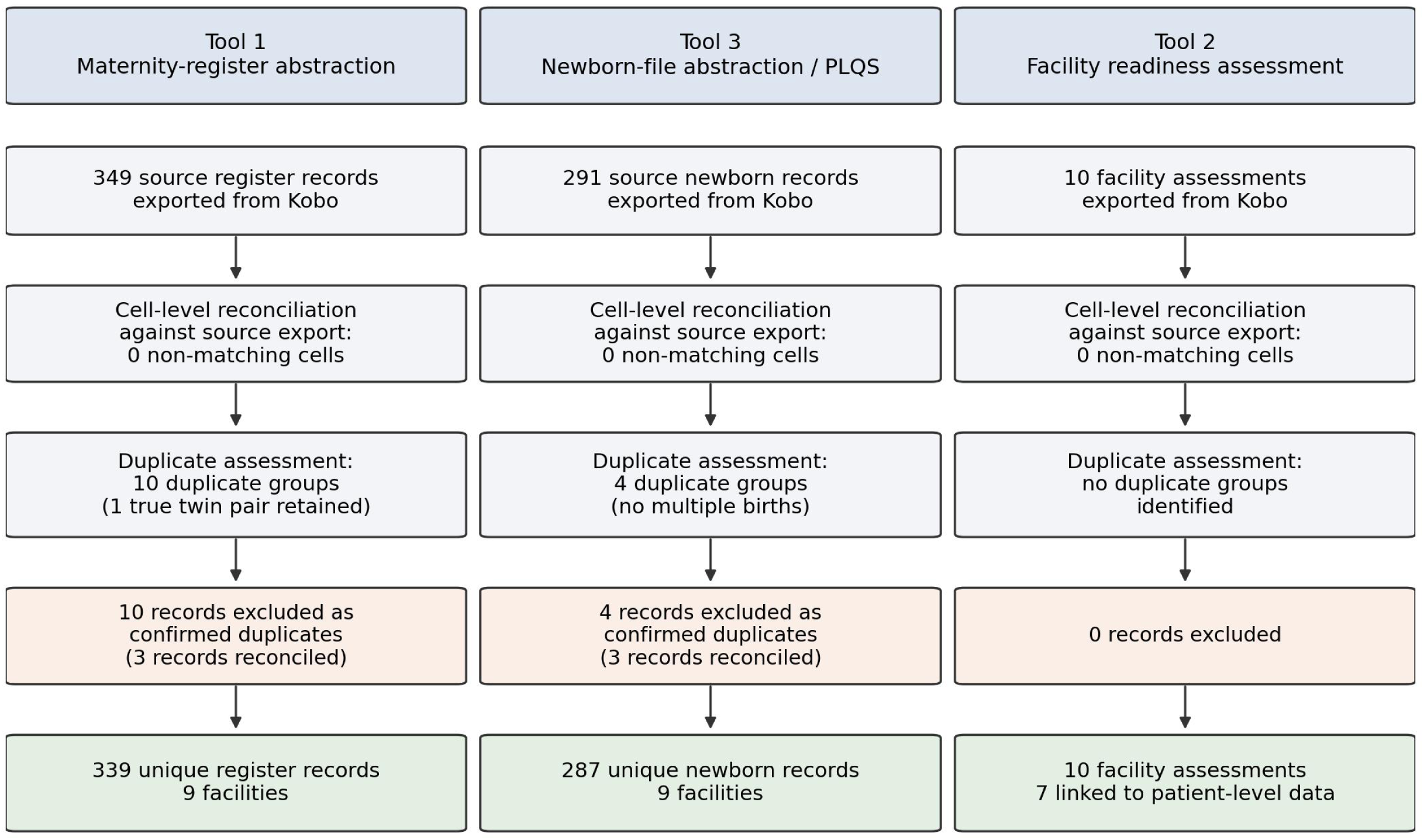
Data-quality assessment flow diagram. Source records, cell-level verification against the original electronic export, duplicate assessment, exclusions and final unique record counts for the maternity-register (Tool 1), newborn clinical record (Tool 3) and facility readiness (Tool 2) datasets. One documented twin pair in the register dataset was retained as two separate newborns and was not treated as a duplicate.

### Completeness of maternity-register data

Register documentation was strong. Facility, register number, date of delivery and birth outcome were documented in all 349 records, and 14 of 19 core variables exceeded 97% completeness (table 2, panel A; online supplemental figure 1; all variables in online supplemental table 2). Median record-level completeness was 100% (IQR 94.7 to 100; mean 96.7%, SD 4.7; range 38.9 to 100), with identification and outcome domains complete for every record. A fully complete core record was nevertheless present in only 175 of 349 (50.1%, 95% CI 44.9 to 55.4), because the shortfall concentrated in three fields most records did not escape: maternal comorbidity status (259/349; 74.2%, 95% CI 69.4 to 78.5, with 52 blanks and 38 explicit ‘not recorded’), antenatal booking trimester (298/349; 85.4%) and residence type (327/349; 93.7%). Gestational age was recorded for 341 records (97.7%) and its source for 343 (98.3%), but the source was last menstrual period in 78.5% and ultrasound in only 19.2%.

**Table 2.**
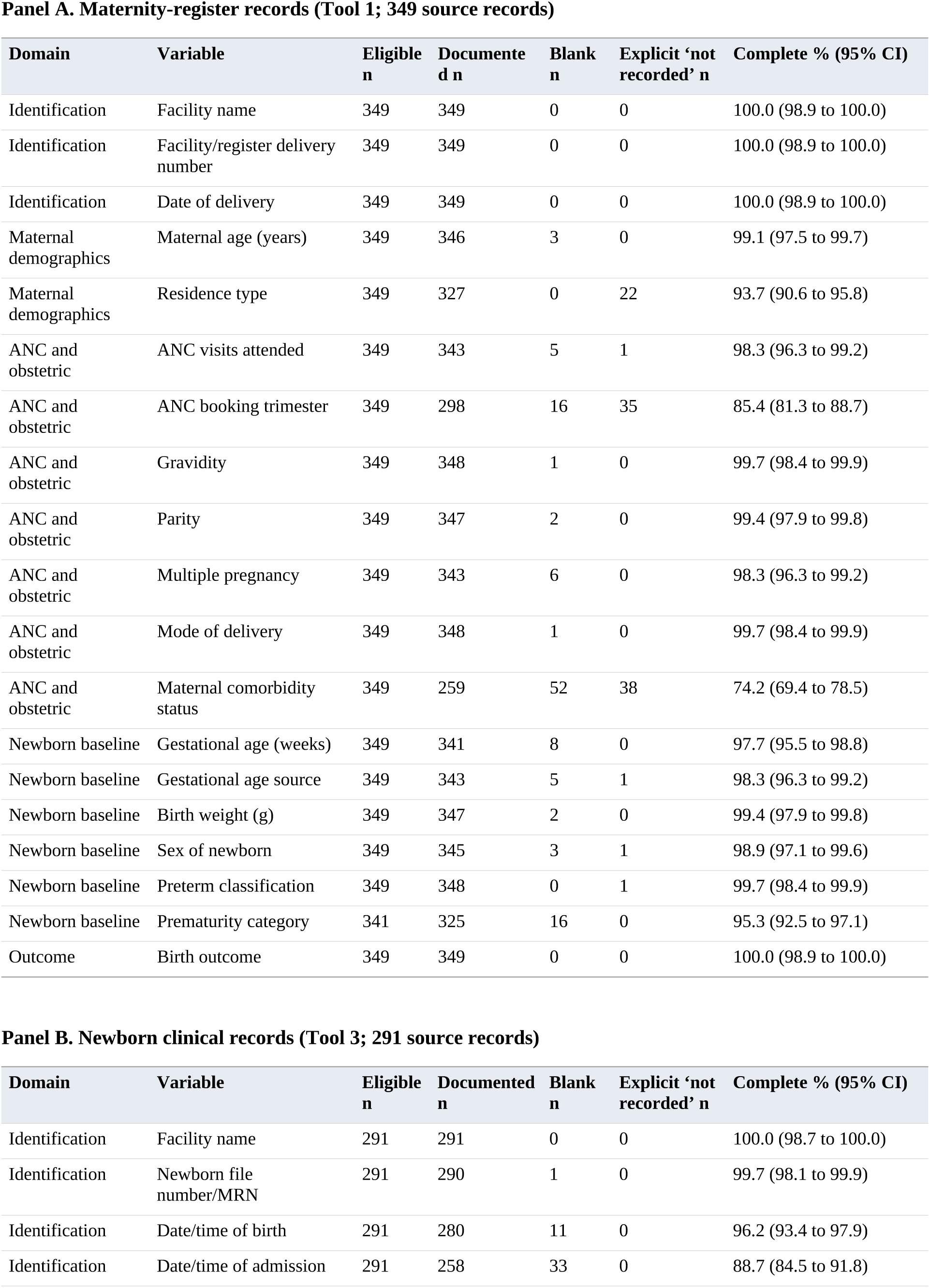
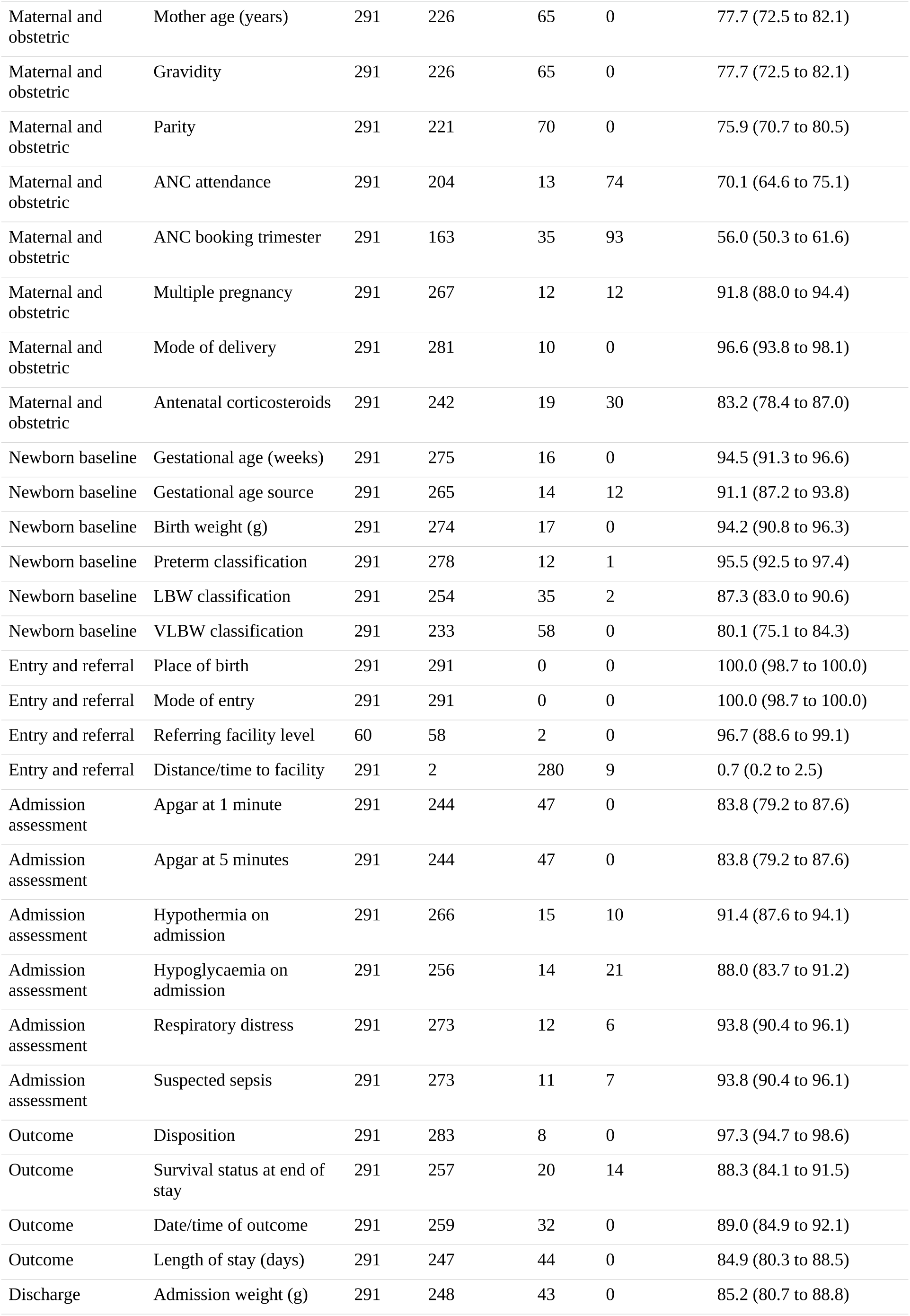
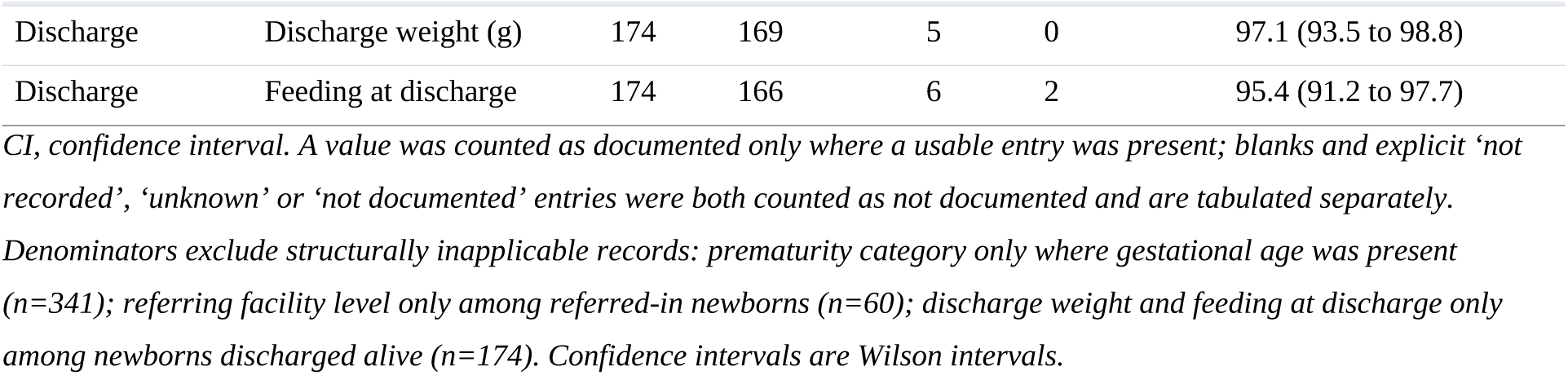
Variable-level documentation completeness in original (uncleaned) abstractions.

### Completeness of newborn clinical records

Newborn records were less complete and far more variable (table 2, panel B). Median record-level completeness was 93.5% (IQR 84.8 to 100; mean 88.5%, SD 17.1; range 12.9 to 100) and a complete core record was present in 77 of 291 records (26.5%, 95% CI 21.7 to 31.8), with domain means ranging from 96.1% for identification to 78.6% for maternal and obstetric information.

Documentation degraded systematically as fields moved away from the newborn’s immediate clinical state. Facility, place of birth and mode of entry were complete in all records and file number in 290/291 (99.7%), but date and time of admission were documented in only 258/291 (88.7%) despite date and time of birth being present in 280/291 (96.2%). Maternal history was weakest: mother’s age and gravidity 226/291 (77.7%) each, parity 221/291 (75.9%), antenatal attendance 204/291 (70.1%) and booking trimester 163/291 (56.0%). Distance or travel time was usable in 2 of 291 records (0.7%, 95% CI 0.2 to 2.5), effectively removing the variable from analysis.

Outcome fields were intermediate: disposition 283/291 (97.3%), survival status 257/291 (88.3%), outcome date and time 259/291 (89.0%), recorded length of stay 247/291 (84.9%) and five-minute Apgar 244/291 (83.8%). Among the 174 newborns discharged alive, discharge weight was documented in 169 (97.1%) and feeding at discharge in 166 (95.4%), so growth and feeding documentation was good where structurally applicable. The 31 PLQS items contained no blank cells; 87.2% of item-cells carried a numerical score and 12.8% were marked not applicable, the kangaroo mother care domain accounting for the largest share (26 to 49 per item).

Two facilities drove much of the variation (figure 2). At one sub-county hospital the entire maternal domain was blank in all 53 records — mother’s age, gravidity, parity and antenatal attendance documented in none — while gestational age, birth weight and disposition were near-complete, a pattern consistent with abstraction from a newborn-unit source that does not carry maternal history. At a second sub-county hospital contributing 16 records, completeness was low across nearly every domain (mean 34.9%, median 14.0%), including no five-minute Apgar and admission date and time in 6 of 16 records.

**Figure 2.**
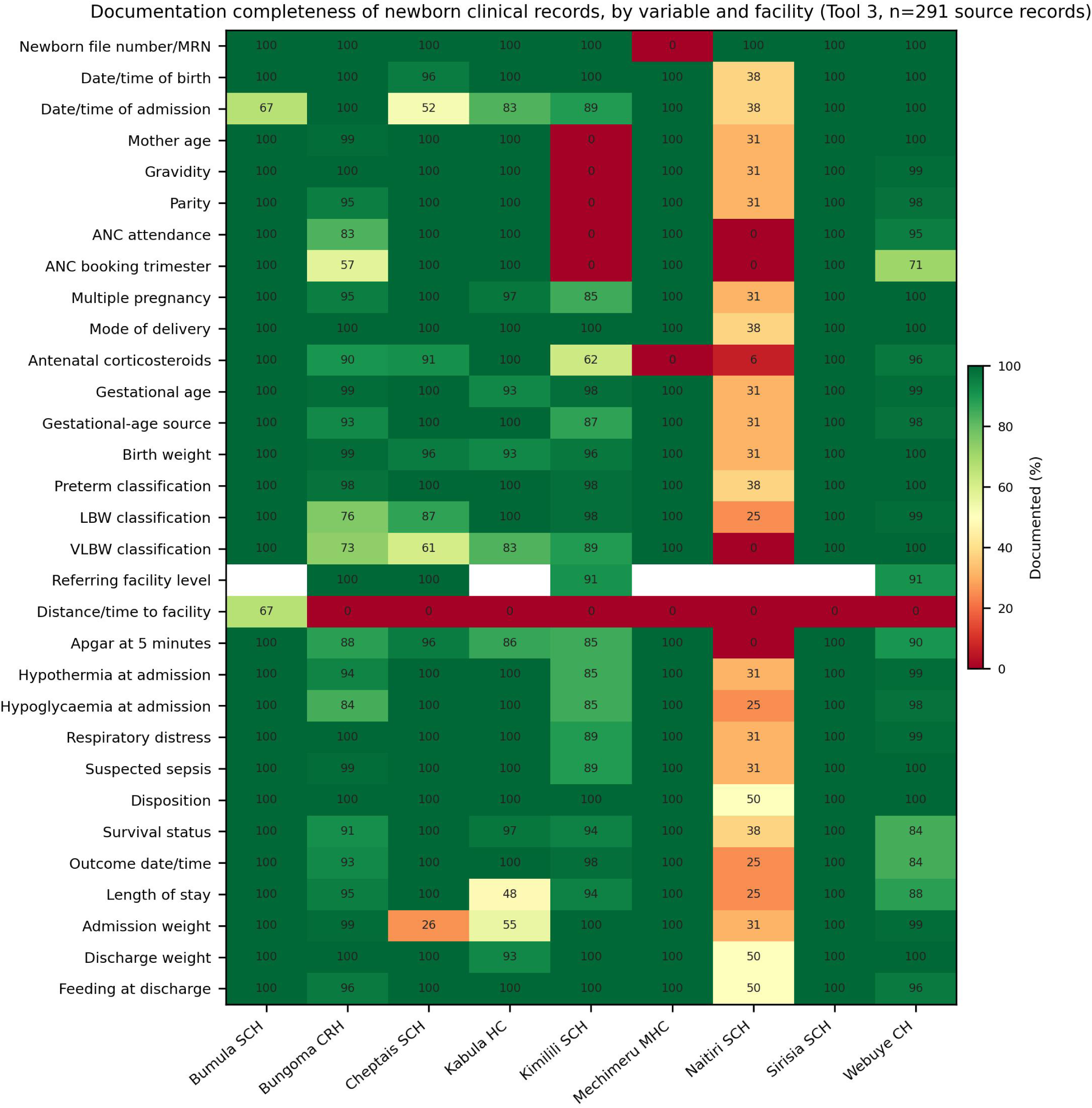
Documentation completeness of newborn clinical records by variable and facility (Tool 3; 291 source records). Cell values are the percentage of that facility’s records in which the variable carried a usable entry, computed from original uncleaned abstractions. Blank cells indicate that no record at that facility was eligible for the variable (referring facility level applies only to referred-in newborns). Facility abbreviations: CRH, county referral hospital; CH, county hospital; SCH, sub-county hospital; HC, health centre; MHC, model health centre.

### Internal consistency and plausibility

Biological plausibility was not the problem: no maternal age, gestational age, birth weight, Apgar score, parity value or daily weight change in either dataset fell outside its prespecified range, and only three gravidity values (0.9%) and two gestational-age and birth-weight combinations (0.6%) were flagged (table 3).

**Table 3.**
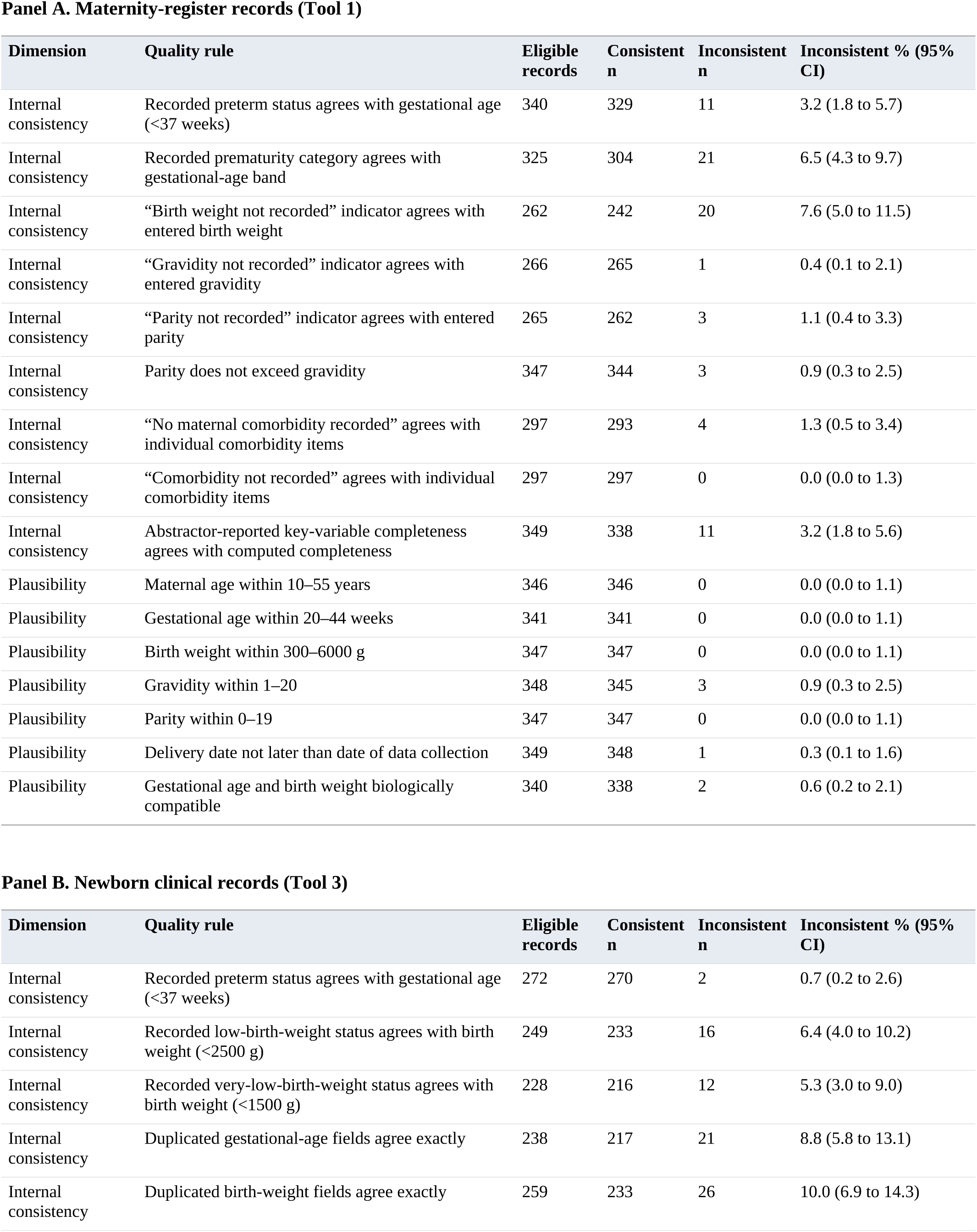
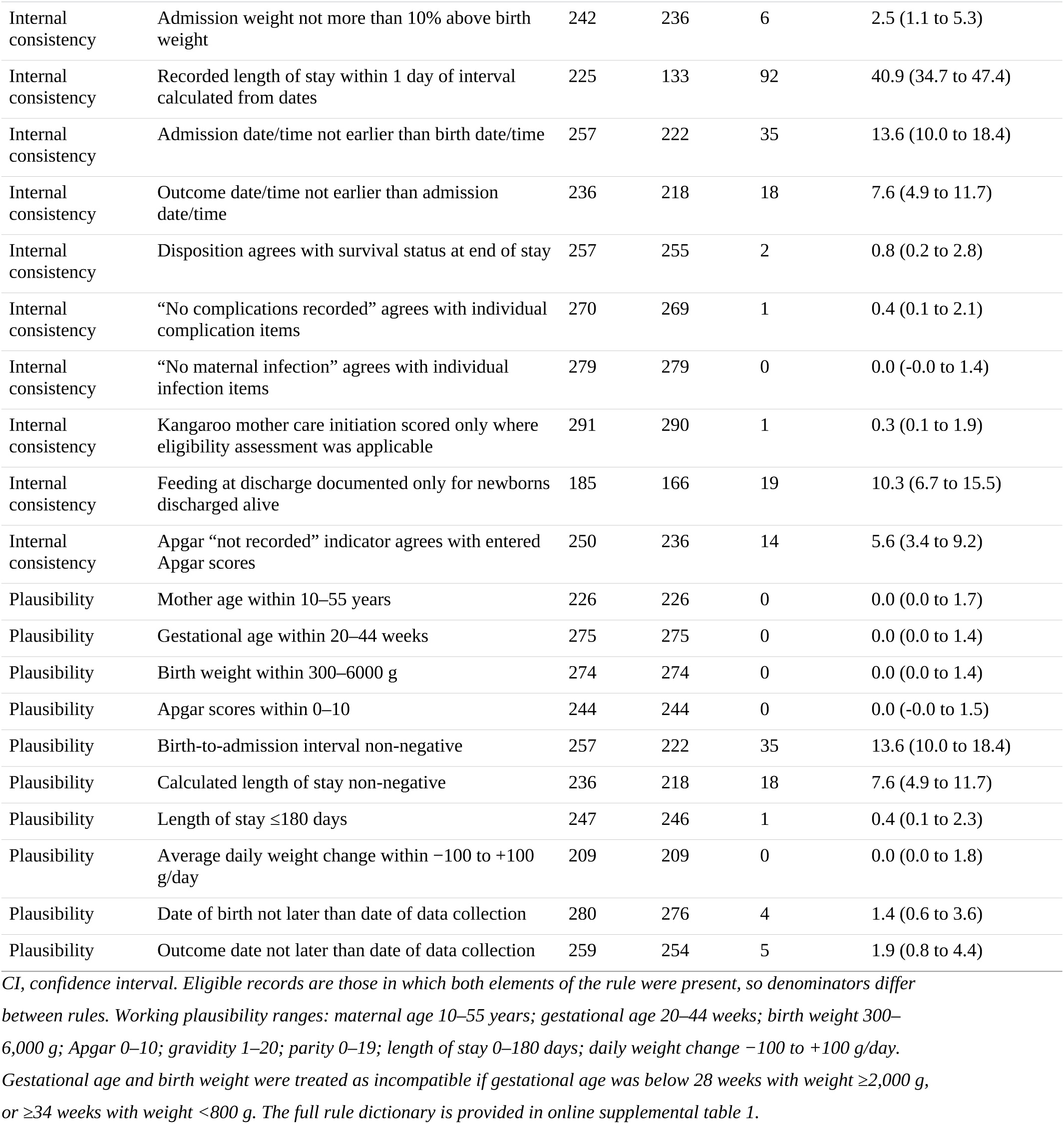
Internal-consistency and plausibility rule violations.

| **Dimension** | **Quality rule** | **Eligible records** | **Consistent n** | **Inconsistent n** | **Inconsistent % (95% CI)** |
|---|---|---|---|---|---|
| **Panel A. Maternity-register records (Tool 1)** |  |  |  |  |  |
| Internal consistency | Recorded preterm status agrees with gestational age (<37 weeks) | 340 | 329 | 11 | 3.2 (1.8 to 5.7) |
| Internal consistency | Recorded prematurity category agrees with gestational-age band | 325 | 304 | 21 | 6.5 (4.3 to 9.7) |
| Internal consistency | “Birth weight not recorded” indicator agrees with entered birth weight | 262 | 242 | 20 | 7.6 (5.0 to 11.5) |
| Internal consistency | “Gravidity not recorded” indicator agrees with entered gravidity | 266 | 265 | 1 | 0.4 (0.1 to 2.1) |
| Internal consistency | “Parity not recorded” indicator agrees with entered parity | 265 | 262 | 3 | 1.1 (0.4 to 3.3) |
| Internal consistency | Parity does not exceed gravidity | 347 | 344 | 3 | 0.9 (0.3 to 2.5) |
| Internal consistency | “No maternal comorbidity recorded” agrees with individual comorbidity items | 297 | 293 | 4 | 1.3 (0.5 to 3.4) |
| Internal consistency | “Comorbidity not recorded” agrees with individual comorbidity items | 297 | 297 | 0 | 0.0 (0.0 to 1.3) |
| Internal consistency | Abstractor-reported key-variable completeness agrees with computed completeness | 349 | 338 | 11 | 3.2 (1.8 to 5.6) |
| Plausibility | Maternal age within 10–55 years | 346 | 346 | 0 | 0.0 (0.0 to 1.1) |
| Plausibility | Gestational age within 20–44 weeks | 341 | 341 | 0 | 0.0 (0.0 to 1.1) |
| Plausibility | Birth weight within 300–6000 g | 347 | 347 | 0 | 0.0 (0.0 to 1.1) |
| Plausibility | Gravidity within 1–20 | 348 | 345 | 3 | 0.9 (0.3 to 2.5) |
| Plausibility | Parity within 0–19 | 347 | 347 | 0 | 0.0 (0.0 to 1.1) |
| Plausibility | Delivery date not later than date of data collection | 349 | 348 | 1 | 0.3 (0.1 to 1.6) |
| Plausibility | Gestational age and birth weight biologically compatible | 340 | 338 | 2 | 0.6 (0.2 to 2.1) |
| **Panel B. Newborn clinical records (Tool 3)** |  |  |  |  |  |
| **Dimension** | **Quality rule** | **Eligible records** | **Consistent n** | **Inconsistent n** | **Inconsistent % (95% CI)** |
| Internal consistency | Recorded preterm status agrees with gestational age (<37 weeks) | 272 | 270 | 2 | 0.7 (0.2 to 2.6) |
| Internal consistency | Recorded low-birth-weight status agrees with birth weight (<2500 g) | 249 | 233 | 16 | 6.4 (4.0 to 10.2) |
| Internal consistency | Recorded very-low-birth-weight status agrees with birth weight (<1500 g) | 228 | 216 | 12 | 5.3 (3.0 to 9.0) |
| Internal consistency | Duplicated gestational-age fields agree exactly | 238 | 217 | 21 | 8.8 (5.8 to 13.1) |
| Internal consistency | Duplicated birth-weight fields agree exactly | 259 | 233 | 26 | 10.0 (6.9 to 14.3) |
| Internal consistency | Admission weight not more than 10% above birth weight | 242 | 236 | 6 | 2.5 (1.1 to 5.3) |
| Internal consistency | Recorded length of stay within 1 day of interval calculated from dates | 225 | 133 | 92 | 40.9 (34.7 to 47.4) |
| Internal consistency | Admission date/time not earlier than birth date/time | 257 | 222 | 35 | 13.6 (10.0 to 18.4) |
| Internal consistency | Outcome date/time not earlier than admission date/time | 236 | 218 | 18 | 7.6 (4.9 to 11.7) |
| Internal consistency | Disposition agrees with survival status at end of stay | 257 | 255 | 2 | 0.8 (0.2 to 2.8) |
| Internal consistency | “No complications recorded” agrees with individual complication items | 270 | 269 | 1 | 0.4 (0.1 to 2.1) |
| Internal consistency | “No maternal infection” agrees with individual infection items | 279 | 279 | 0 | 0.0 (-0.0 to 1.4) |
| Internal consistency | Kangaroo mother care initiation scored only where eligibility assessment was applicable | 291 | 290 | 1 | 0.3 (0.1 to 1.9) |
| Internal consistency | Feeding at discharge documented only for newborns discharged alive | 185 | 166 | 19 | 10.3 (6.7 to 15.5) |
| Internal consistency | Apgar “not recorded” indicator agrees with entered Apgar scores | 250 | 236 | 14 | 5.6 (3.4 to 9.2) |
| Plausibility | Mother age within 10–55 years | 226 | 226 | 0 | 0.0 (0.0 to 1.7) |
| Plausibility | Gestational age within 20–44 weeks | 275 | 275 | 0 | 0.0 (0.0 to 1.4) |
| Plausibility | Birth weight within 300–6000 g | 274 | 274 | 0 | 0.0 (0.0 to 1.4) |
| Plausibility | Apgar scores within 0–10 | 244 | 244 | 0 | 0.0 (-0.0 to 1.5) |
| Plausibility | Birth-to-admission interval non-negative | 257 | 222 | 35 | 13.6 (10.0 to 18.4) |
| Plausibility | Calculated length of stay non-negative | 236 | 218 | 18 | 7.6 (4.9 to 11.7) |
| Plausibility | Length of stay ≤180 days | 247 | 246 | 1 | 0.4 (0.1 to 2.3) |
| Plausibility | Average daily weight change within −100 to +100 g/day | 209 | 209 | 0 | 0.0 (0.0 to 1.8) |
| Plausibility | Date of birth not later than date of data collection | 280 | 276 | 4 | 1.4 (0.6 to 3.6) |
| Plausibility | Outcome date not later than date of data collection | 259 | 254 | 5 | 1.9 (0.8 to 4.4) |

*CI, confidence interval. Eligible records are those in which both elements of the rule were present, so denominators differ between rules. Working plausibility ranges: maternal age 10–55 years; gestational age 20–44 weeks; birth weight 300–6,000 g; Apgar 0–10; gravidity 1–20; parity 0–19; length of stay 0–180 days; daily weight change −100 to +100 g/day. Gestational age and birth weight were treated as incompatible if gestational age was below 28 weeks with weight ≥2,000 g, or ≥34 weeks with weight <800 g. The full rule dictionary is provided in online supplemental table 1.*

Chronology was. Recorded length of stay differed from the date-derived interval by more than one day in 92 of 225 newborn records (40.9%, 95% CI 34.7 to 47.4); admission preceded birth in 35 of 257 records with both timestamps (13.6%, 95% CI 10.0 to 18.4), of which 25 by more than one hour and 14 by more than 30 days, indicating year or month transcription errors rather than clock imprecision; and the outcome date-time preceded admission in 18 of 236 (7.6%). Four birth dates and five outcome dates in newborn records, and one delivery date in the register, were later than the date on which the record was abstracted, one birth date by more than nine months.

Contradictions clustered around fields captured more than once or alongside a ‘not recorded’ switch. The two birth-weight fields disagreed in 26 of 259 paired records (10.0%) and the two gestational-age fields in 21 of 238 (8.8%). Recorded LBW status contradicted the entered birth weight in 16 of 249 records (6.4%) and VLBW status in 12 of 228 (5.3%). Feeding at discharge was documented for 19 newborns not discharged alive (10.3% of completed fields). In the register dataset, the ‘birth weight not recorded’ switch contradicted the entered weight in 20 of 262 records (7.6%), the recorded prematurity category disagreed with the gestational-age band in 21 of 325 (6.5%), and the abstractor’s assessment that key variables were complete disagreed with computed completeness in 11 of 349 (3.2%). Composite ‘none recorded’ indicators performed well, with no maternal-infection contradiction and only four comorbidity contradictions (1.3%).

### Agreement between recorded and rule-derived classifications

Agreement varied by classification and was consistently limited by the rarity of the negative category (table 4). In the register dataset, recorded preterm status agreed with the gestational-age-derived classification in 329 of 340 records (96.8%), but because 339 of 349 records were classified preterm, kappa was only 0.40 (95% CI 0.12 to 0.69) with negative agreement of 42.1%. The multi-category prematurity classification performed better, with 93.5% agreement and kappa 0.87 (95% CI 0.82 to 0.93); disagreements concentrated at the 28-week boundary, where positive agreement for extremely preterm was 69.6% against 98.1% for moderate-to-late preterm.

**Table 4.**
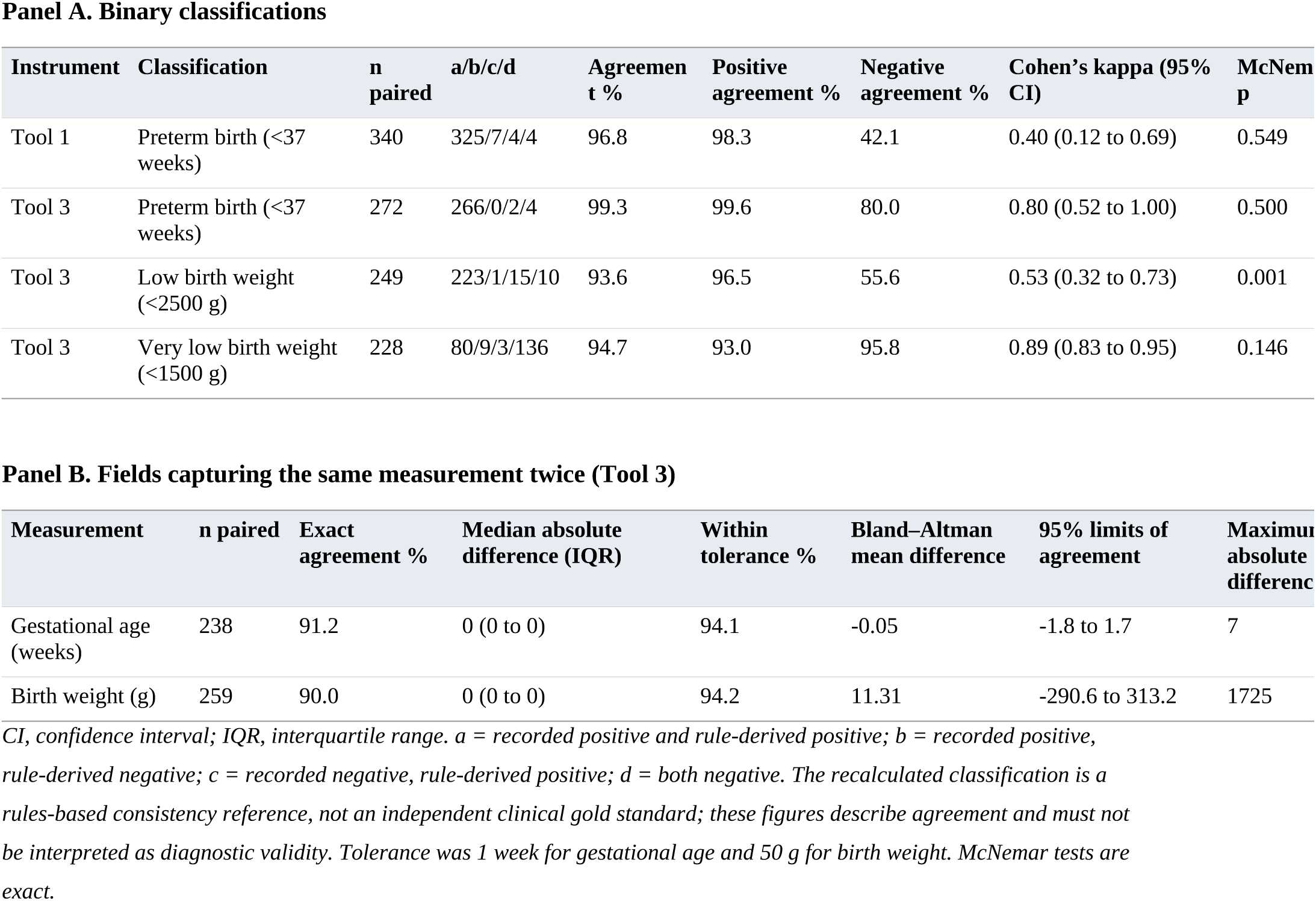
Agreement between recorded classifications and classifications recalculated from the underlying measurement.

In the newborn dataset, preterm status agreed in 266 of 272 records (99.3%; kappa 0.80, 95% CI 0.52 to 1.00) and VLBW status in 216 of 228 (94.7%; kappa 0.89, 95% CI 0.83 to 0.95). LBW classification was weakest: 233 of 249 agreed (93.6%) but kappa was 0.53 (95% CI 0.32 to 0.73), and disagreement was directional — 15 newborns with a birth weight below 2,500 g were not recorded as LBW while only one was recorded as LBW without a qualifying weight (exact McNemar p=0.001) — indicating systematic under-recording rather than random error.

Where the same measurement appeared in two fields, most pairs agreed exactly (gestational age 91.2%, birth weight 90.0%) and most of the remainder fell within tolerance (94.1% within 1 week; 94.2% within 50 g). The disagreements that remained were large rather than marginal: Bland–Altman limits of agreement for birth weight were −291 g to +313 g with a maximum absolute difference of 1,725 g, and the maximum gestational-age difference was 7 weeks (online supplemental figure 2). A small minority of records therefore contain a discrepancy wide enough to move a newborn between weight categories.

### Analytical usability

Table 5 summarises what the datasets can and cannot support. Preterm identification, prematurity severity, birth-weight classification, stillbirth among recorded deliveries, in-hospital mortality among newborns with a known final outcome, and referral status were fully supported or supported with limitations. Length of stay was computable from valid dates in 218 of 291 records (74.9%) and weight change from paired admission and discharge weights in 219 (75.3%); a known final outcome was available for 239 of 291 (82.1%), with 44 referred out or still admitted and 8 with no disposition recorded.

**Table 5.** Fitness-for-analysis matrix: analyses the routine data can and cannot support.

| Analysis | Assessment | Basis and reason |
|---|---|---|
| Identification of preterm status | Fully supported | Gestational age documented in 97.7% of register and 94.5% of newborn records; recorded and rule-derived classification agreed in 96.8% and 99.3% respectively. |
| Prematurity severity (three categories) | Fully supported | Recorded category agreed with the gestational-age band in 93.5% of register records (kappa 0.87); disagreements concentrated at the 28-week boundary and should be resolved from source where extremely preterm status is the outcome. |
| Low-birth-weight classification | Supported with limitations | Birth weight documented in 99.4% of register and 94.2% of newborn records, but the recorded LBW flag under-identified 15 of 238 qualifying newborns; analyses should derive the flag from the recorded weight rather than use the recorded flag. |
| Very-low-birth-weight classification | Fully supported | Agreement with the weight-derived classification was 94.7% (kappa 0.89). |
| Stillbirth among recorded deliveries | Fully supported | Birth outcome documented in all 349 register records, with fresh and macerated stillbirth distinguished. |
| In-hospital mortality | Supported with limitations | Restricted to the 239 of 291 newborns (82.1%) with a known final outcome; 25 referred out and 19 still admitted have no ascertained endpoint and must not be counted as survivors. |
| Referral status (mode of entry, place of birth) | Fully supported | Both documented in all 291 newborn records. |
| Referral timing | Supported with limitations | A birth-to-admission interval was computable and non-negative in 222 of 291 records (76.3%); it describes an interval, not a transport process. |
| Transport process (mode, oxygen, escort, departure time) | Not currently supported | Distance or travel time was usable in 2 of 291 records (0.7%) and no structured transport fields were captured. |
| Length of stay | Supported with limitations | Computable from valid dates in 218 of 291 records (74.9%); recorded and date-derived values differed by more than one day in 40.9% of records with both, so the two are not interchangeable. |
| Weight change and average daily weight gain | Supported with limitations | Paired admission and discharge weights available for 219 of 291 records (75.3%); restricted to discharged survivors and therefore subject to survivor selection. |
| Feeding at discharge | Supported with limitations | Documented for 166 of 174 newborns discharged alive (95.4%), but was also completed for 19 newborns not discharged alive, which must be treated as missing. |
| Patient-level quality score (PLQS) | Fully supported | No blank item cells; 87.2% of item-cells carried a numerical score, with 12.8% marked not applicable and excluded from denominators. |
| Kangaroo mother care implementation | Supported with limitations | Not-applicable responses ranged from 26 to 49 per KMC item, so eligibility and duration denominators are incompletely defined. |
| Facility-level comparison of documentation | Supported with limitations | Nine facilities contribute patient-level records and seven have linked readiness data; comparisons are descriptive and ecological. |
| Preterm-birth prevalence | Not currently supported | No facility-period total-delivery denominator was captured and 339 of 349 register records were already classified preterm; recovery of facility-month totals for all deliveries, live births and stillbirths would make estimation possible. |

*LBW, low birth weight; PLQS, patient-level quality score; KMC, kangaroo mother care. Percentages are computed on original (uncleaned) abstractions.*

Two analyses were not supported. Referral timing was computable in 222 of 291 records (76.3%) once invalid intervals were excluded, but structured transport variables — departure and arrival time, mode, oxygen and escort — were absent, so referral delay can be described only as a birth-to-admission interval, not as a transport process. Preterm-birth prevalence could not be estimated at all: the register export contains no facility-period total-delivery denominator and 339 of 349 abstracted records were already classified preterm, so dividing recorded preterm births by abstracted records would return a figure describing the abstraction rather than the population. Prevalence estimation requires facility-month totals for all deliveries, live births and stillbirths, which were not collected.

### Facility variation and information-system readiness

Mean register completeness ranged from 93.1% to 99.3% across the nine facilities, but the proportion of complete core records ranged from 0% to 86.7% and the record-level inconsistency rate from 2.1% to 70.6%; newborn-record completeness varied far more widely, from 34.9% to 100% (online supplemental figure 3; online supplemental table 3).

Recalculated HIS readiness ranged from 50.0% to 100.0% and QI and governance readiness from 12.5% to 100.0%, with six of ten facilities scoring 12.5% on the QI domain, reflecting an absent QI team and no review meetings. Among the seven facilities contributing both readiness and patient-level data, correlations were positive but imprecise and none reached conventional significance: overall readiness with the proportion of complete core newborn records gave rho 0.66 (95% CI −0.25 to 0.95, p=0.111); HIS readiness with mean register completeness rho 0.27 (95% CI −0.64 to 0.87, p=0.554); and HIS readiness with mean newborn-record completeness rho 0.04 (95% CI −0.76 to 0.79, p=0.938) (online supplemental figure 4). With seven facilities these relationships are ecological, exploratory and compatible with a wide range of underlying associations.

### Factors associated with complete documentation

Predictors differed sharply between the two record types (table 6). For register records, only facility level was associated with complete documentation: level 5 versus level 4 gave an adjusted prevalence ratio of 2.24 (95% CI 1.82 to 2.75), with no association for period of delivery or birth outcome. In the model restricted to facilities with a readiness assessment (n=279 records, 140 complete), each 25-percentage-point increment in HIS readiness was associated with a prevalence ratio of 1.44 (95% CI 1.04 to 1.99), while QI readiness showed no association (0.93, 95% CI 0.82 to 1.05).

**Table 6.** Factors associated with a complete core record (modified Poisson regression with robust variance)

| Instrument and factor | Records | Complete, n (%) | Crude PR (95% CI) | Adjusted PR (95% CI) | p value |
|---|---|---|---|---|---|
| **Panel A. Maternity register (Tool 1); n=349, 175 complete (50.1%)** |  |  |  |  |  |
| Facility level 4 (reference) | 225 | 88 (39.1) | 1.00 | 1.00 |  |
| Facility level 3 | 48 | 23 (47.9) | 1.23 (0.88 to 1.72) | 1.17 (0.82 to 1.69) | 0.386 |
| Facility level 5 | 76 | 64 (84.2) | 2.15 (1.78 to 2.60) | 2.24 (1.82 to 2.75) | <0.001 |
| Delivery Jul 2024–Dec 2025 (reference) | 77 | 35 (45.5) | 1.00 | 1.00 |  |
| Delivery Jan–May 2026 | 272 | 140 (51.5) | 1.13 (0.86 to 1.48) | 0.89 (0.64 to 1.23) | 0.470 |
| Live birth (reference) | 300 | 151 (50.3) | 1.00 | 1.00 |  |
| Stillbirth | 49 | 24 (49.0) | 0.97 (0.72 to 1.32) | 0.87 (0.66 to 1.14) | 0.303 |
| **Panel B. Maternity register, facilities with a readiness assessment; n=279, 140 complete** |  |  |  |  |  |
| HIS readiness, per 25 percentage points | 279 | – | – | 1.44 (1.04 to 1.99) | 0.028 |
| QI and governance readiness, per 25 percentage points | 279 | – | – | 0.93 (0.82 to 1.05) | 0.249 |
| **Panel C. Newborn clinical record (Tool 3); n=291, 77 complete (26.5%)** |  |  |  |  |  |
| Facility level 4 (reference) | 178 | 48 (27.0) | 1.00 | 1.00 |  |
| Facility level 3 | 30 | 7 (23.3) | 0.87 (0.43 to 1.73) | 2.36 (0.95 to 5.84) | 0.064 |
| Facility level 5 | 83 | 22 (26.5) | 0.98 (0.64 to 1.51) | 0.49 (0.26 to 0.92) | 0.027 |
| Newborn unit (reference) | 218 | 63 (28.9) | 1.00 | 1.00 |  |
| Maternity, KMC or other ward | 73 | 14 (19.2) | 0.66 (0.40 to 1.11) | 0.60 (0.30 to 1.21) | 0.154 |
| Fewer than three record sources (reference) | 189 | 46 (24.3) | 1.00 | 1.00 |  |
| Three or more record sources | 102 | 31 (30.4) | 1.25 (0.85 to 1.84) | 1.80 (1.05 to 3.08) | 0.032 |
| Direct admission (reference) | 231 | 63 (27.3) | 1.00 | 1.00 |  |
| Referred in | 60 | 14 (23.3) | 0.86 (0.52 to 1.42) | 0.88 (0.52 to 1.48) | 0.619 |
| Disposition other than died (reference) | 226 | 67 (29.6) | 1.00 | 1.00 |  |
| Died before discharge | 65 | 10 (15.4) | 0.52 (0.28 to 0.95) | 0.46 (0.25 to 0.84) | 0.011 |
| Birth before Jan 2026 (reference) | 74 | 7 (9.5) | 1.00 | 1.00 |  |
| Birth from Jan 2026 onwards | 217 | 70 (32.3) | 3.41 (1.64 to 7.08) | 4.43 (2.29 to 8.57) | <0.001 |
| **Panel D. Newborn clinical record, facilities with a readiness assessment; n=215, 75 complete** |  |  |  |  |  |
| HIS readiness, per 25 percentage points | 215 | – | – | 1.32 (0.74 to 2.35) | 0.353 |
| QI and governance readiness, per 25 percentage points | 215 | – | – | 1.39 (0.99 to 1.94) | 0.056 |

*PR, prevalence ratio; CI, confidence interval; HIS, health information system; QI, quality improvement; KMC, kangaroo mother care. Panel A adjusts for all factors shown in that panel; panel C adjusts for all factors shown in that panel. Panels B and D are mutually adjusted for the two readiness scores and are restricted to the seven facilities with a Tool 2 assessment. p values are for the adjusted estimate. A complete core record required documentation of every applicable core field (19 register fields; 34 newborn fields, excluding distance or travel time).*

For newborn records, the strongest association was temporal: records for newborns born from January 2026 onwards were markedly more likely to be complete (adjusted prevalence ratio 4.43, 95% CI 2.29 to 8.57). Consulting three or more record sources was associated with more complete abstraction (1.80, 95% CI 1.05 to 3.08), and records for newborns who died were less complete (0.46, 95% CI 0.25 to 0.84), consistent with reduced opportunity to document care and outcome for newborns who die early. Level 5 records were less complete than level 4 records after adjustment (0.49, 95% CI 0.26 to 0.92), the opposite of the register finding — the referral hospital documents registers well but carries the highest patient volume and the greatest reliance on parallel clinical sources. Ward or unit and mode of entry were not associated with completeness.

Sensitivity analyses (online supplemental table 5) did not alter these conclusions. Restricting to unique records after duplicate resolution changed mean register completeness by less than 0.1 percentage points and newborn completeness by 0.2. Counting any entry, including explicit ‘not recorded’, as documented raised mean completeness to 99.1% and 90.3% respectively, showing that much of the newborn shortfall is genuine blank space. Restricting to facilities contributing at least ten records, and excluding records with invalid date sequences, produced estimates within 1.4 percentage points of the primary analysis.

### Provider perspectives

Both providers described parallel paper and electronic pathways. Structured paper forms are transcribed after discharge — ‘As we take care of the baby, after discharging the baby, they take the patient file and enter that information’ (Provider KII 1) — and the unit receives monthly Clinical Information Network reports and partner dashboards, using both: ‘This data, we use it to make decisions.’ Provider KII 2 described the paper layer: ‘we have the daily bed return. We have the admission registers. Then we have the kangaroo mother care register.’

Both located the failure point at the moment of recording rather than in the tools. Provider KII 2 stated it directly: ‘in terms of documentation, yes, we have the registers and the papers required for documentation. But now, we normally miss each other. We normally focus on, is this baby sick? Yes. Is the baby out of danger? Yes.’ The same provider linked monitoring documentation explicitly to staffing: ‘we also monitor vitals hourly. That is the bad thing. Hourly with the shortage, sometimes we miss, sometimes we do it.’

Audit and feedback were described as established and as having changed in character: ‘At first, we thought that audits were for shame, but as we’ve been carrying them out, they’ve been educational for us’ (Provider KII 2). Unit reviews are weekly, data are audited monthly, and errors identified centrally are returned for clarification: ‘when that data is having errors, I’ll be called to clarify what happened on such and such a baby, or such and such a file.’ Perinatal death review was described as durable — ‘every third Friday of the month is MPDSR. So it’s permanent’ (Provider KII 1) — but implementation of resulting recommendations was the identified weak link: ‘Our only weakness has been in implementation of actual plans because some gaps keep recurring and recurring.’ Workload displaced scheduled review: ‘we had like 18 admissions and we are supposed to have a meeting at 10, we postponed it, we didn’t do it.’ Free-text facility responses converged on the same constraint, staff shortage being named as a major gap by nine of ten facilities.

### Integrated mixed-method findings

The joint display is presented in online supplemental table 6. Integration was convergent for the staffing–completeness relationship, where the quantitative gradient in maternal-history and monitoring fields matched the providers’ account of documentation deferred under clinical pressure, and for the mortality gradient in documentation completeness. It was complementary for the electronic-system findings: the presence of CIN and dashboard reporting explains why the referral hospital’s register documentation and identification fields perform well without explaining why its individual newborn records do not. It was expansive for duplicated fields and free-text facility naming, which providers did not raise but which the quantitative analysis identified as material threats to linkage. It was discordant in one respect: providers described monthly data review as routine and corrective, yet the chronological-error rate suggests date-sequence validation is not part of what is reviewed.

## Discussion

### Principal findings

Across twelve facilities in Western Kenya, routine maternity and newborn records were substantially complete for the fields that identify a newborn and describe their immediate clinical state, and substantially incomplete for the fields describing when things happened, what preceded the birth, and how the newborn arrived. Median record completeness was 100% for register records and 93.5% for newborn records, but only 26.5% of newborn records had every applicable core field documented. Biological implausibility was essentially absent; chronological error was not, with over 40% of newborn records showing a length-of-stay discrepancy exceeding one day and 13.6% recording admission before birth. Recorded classifications agreed well with recalculated classifications for VLBW but only moderately for LBW, with directional under-recording. Preterm-birth prevalence could not be estimated because no all-delivery denominator exists.

### Interpreting the completeness findings

The register findings sit at the better end of published experience. The systematic review of routine facility data for newborn indicators in low-and middle-income countries found register completeness above 60% for gestational age, sex, birth weight and place of delivery but with wide ranges,[10] and a data-quality strengthening campaign across maternity registers in Kenya and Uganda reported centre-averaged completeness of 87–97% for birth weight and 52–92% for gestational age.[31] Our register gestational-age completeness of 97.7% and birth-weight completeness of 99.4% sit at or above the top of those ranges. The Kenyan national KHIS evaluation reported a mean facility-quarter completeness score of 61.1% for aggregate reporting,[12] and the gap is instructive: paper registers are the first point of capture and are typically more complete than the aggregate summaries derived from them, so completeness measured at the register overstates what reaches the national system.

The newborn-record findings are more concerning precisely because these are the records clinical audit and quality improvement depend on. The pattern is not random attrition: it follows a gradient from the newborn’s present state (near-complete) through the newborn’s outcome (85–97%) to the mother’s history (56–78%) to the transport process (0.7%). This is what one would expect if documentation is completed at the bedside under time pressure, prioritising information needed for the next clinical decision — the mechanism providers in the EN-BIRTH register study described when asked why fields go unrecorded.[32] Provider KII 2’s formulation, that the focus is whether the baby is sick and whether the baby is out of danger, describes exactly this gradient. The facility-level outliers reinforce the point: at one hospital an entire maternal domain was blank in all 53 records while newborn clinical fields were near-complete, which is a source-document problem rather than a diligence problem.

### Consistency, classification agreement and their consequences

The directional under-recording of LBW status is the most clinically consequential consistency finding. Fifteen newborns weighing under 2,500 g were not flagged as LBW while only one was flagged without a qualifying weight. If the recorded flag rather than the measured weight drives triage, referral or reporting, a subset of low-birth-weight newborns becomes invisible. Comparable directional discordance has been reported elsewhere: the EN-BIRTH validation study found that register-recorded coverage systematically diverged from observed coverage for several newborn interventions, and that accuracy varied between hospitals even where register design was identical.[33,34] Our finding differs in an important way — we compared a recorded flag against a measurement written in the same record, so the discordance cannot be attributed to differences between data sources. Multi-country record review has likewise found that the presence of a field in an instrument is a poor guide to whether it is completed usefully.[35,36]

The chronological findings deserve emphasis because they are rarely reported. Most published assessments of newborn records address completeness and, less often, internal consistency of values; date-sequence validity is seldom examined.[10,37] Yet the fields affected are precisely those needed to describe time-critical care. An admission timestamp preceding birth by more than 30 days is a transcription error that no plausibility range on a clinical value would detect, and a length-of-stay discrepancy above one day in 40.9% of records means that recorded and date-derived length of stay are not interchangeable. For a county wishing to monitor birth-to-admission intervals as a referral-quality indicator, this is the binding constraint.

The duplicated-field findings point to an instrument design lesson. Capturing gestational age and birth weight twice created a useful internal check, and roughly 90% of pairs agreed exactly, but the disagreements that occurred were wide — up to 1,725 g and 7 weeks. Repeated capture is valuable for detecting error, but only if the discrepancies it surfaces are resolved rather than carried forward.

### The denominator problem

That preterm-birth prevalence cannot be estimated is a finding rather than a failure of analysis, and it generalises beyond this study. The register instrument captured individual delivery records and a preterm classification but no facility-period total-deliveries field, and the realised abstraction was overwhelmingly preterm. This is the structural issue identified in the global assessment of national routine data for low birthweight and preterm births, which found that gaps in gestational-age metadata rather than in birthweight capture are the principal obstacle to routine preterm estimation in sub-Saharan Africa and South Asia,[6] and work in Western Kenya on using maternity-register gestational age reached a similar conclusion about what registers can bear.[7] A numerator-only extract is easy to produce and difficult to detect once analysis begins. Recovering facility-month totals for all deliveries, live births and stillbirths would make prevalence estimable without any change to the individual-record instrument.

### Facility variation and information-system readiness

Facility variation in newborn-record completeness (34.9% to 100%) far exceeded variation in register completeness (93.1% to 99.3%), suggesting the constraint is local documentation practice and source-document availability rather than instrument design. The association between HIS readiness and register completeness (adjusted prevalence ratio 1.44 per 25 percentage points) was in the expected direction, but facility-level correlations were imprecise and non-significant with seven facilities and should not be read as evidence of either presence or absence of an effect; multi-facility assessments in Tanzania and Ethiopia suggest facility-level determinants become estimable only at substantially larger scale.[37,38] The finding that level 5 records were more complete in the register but less complete at the newborn level after adjustment is worth noting for planners: it is compatible with a referral hospital that has invested in register systems and electronic reporting while carrying the highest volume and the most fragmented set of bedside sources, and cautions against assuming that information-system investment at higher levels automatically produces better individual clinical documentation. The markedly higher completeness of records for newborns born from January 2026 may reflect the condition of files at retrieval, recent files being more often intact and in active use, rather than a change in documentation practice, and should not be read as a trend.

### Provider explanations and the audit paradox

The qualitative component identified an implementation gap the quantitative data alone would not have revealed. Both providers described functioning audit structures — weekly unit reviews, monthly data audits, a fixed monthly perinatal death review — and described audit as having shifted from a blaming exercise to an educational one, precisely the culture change that assessments of maternal and perinatal death surveillance and response across sub-Saharan Africa identify as necessary and often absent.[39,40] Yet Provider KII 1 named implementation of action plans as the standing weakness, and Provider KII 2 described a scheduled review postponed because of an 18-admission day. Analyses of MPDSR across African countries have repeatedly found that reports can appear comprehensive while masking weak follow-through on recommendations,[39] and our data are consistent with that pattern in a specific and testable way: date-sequence errors persist at high frequency in a setting where monthly data review demonstrably occurs, which implies that what is reviewed does not include the checks that would catch them.

This suggests a targeted rather than a general remedy. The problem is not the absence of audit but the absence of specific validation rules within it. Automated date-sequence and range checks at the point of data entry would convert a class of error that currently survives monthly review into one that cannot be entered at all. Before-and-after work in Nigerian primary facilities showed that targeted register redesign and supervision improved routine maternal and newborn data quality,[41,42] the Kenyan Clinical Information Network experience shows documentation completeness improving measurably after hospitals join a structured audit-and-feedback network,[15,16,17] as does the Kenyan experience with repeated electronic data quality assessment,[43] and improvements in admission-temperature documentation within the same network illustrate what is achievable for a single field.[44]

## Strengths

The principal methodological strength is that every estimate was computed from the original, unedited abstractions; had cleaned or imputed values been used, completeness would have been overstated. Second, every dataset was verified cell by cell against the original electronic export before analysis, so the estimates are traceable to the source data. Third, prespecified rules were applied across three linked data sources rather than a single register, allowing consistency to be assessed within and between instruments. Fourth, distinguishing blanks from explicit ‘not recorded’ responses showed that most of the newborn shortfall is genuine blank space, which has different implications for remediation.

## Limitations

Ten limitations bear on interpretation. First, no independent clinical gold standard was collected, so accuracy was not and cannot be assessed here; recalculated classifications served only as a rules-based consistency reference. Second, documentation quality is not care quality — a missing field does not establish that care was not delivered, and a complete field does not establish that it was delivered well. Third, missing documentation is not evidence of omitted care, particularly for newborns who died early, whose records were measurably less complete. Fourth, the register dataset lacks a valid all-delivery denominator, so no prevalence claim can be supported. Fifth, only ten facilities contributed readiness data and seven contributed both readiness and patient-level records, so facility-level relationships are exploratory and imprecise. Sixth, the qualitative component comprises two interviews from a single referral hospital and cannot represent documentation practice at lower-level facilities, where the quantitative data suggest the problems differ in kind. Seventh, retrospective abstraction introduces its own error, and abstractor-reported completeness disagreed with computed completeness in 3.2% of register records. Eighth, duplicate records required reconciliation; although exclusions are reversible and conflicts preserved, some contradictory values remain unresolved because source documents could not be re-consulted. Ninth, facility-level correlation cannot establish causality, and the association between HIS readiness and completeness is equally compatible with better-resourced facilities performing better on both. Tenth, one facility lies outside Bungoma County; it contributed readiness data only and no patient-level record, so it does not affect any completeness estimate.

## Implications for practice, policy and research

Four changes would address most of what we found, and none requires new instruments. Making admission date and time, survival status and outcome date and time mandatory fields with point-of-entry date-sequence validation would remove the largest single class of error. Adding facility-month totals for all deliveries, live births and stillbirths to routine reporting would make preterm and low-birth-weight prevalence estimable from data already being collected. Deriving the LBW and VLBW flags automatically from the recorded weight, rather than capturing them as independent items, would eliminate the directional discordance observed. Assigning KHIS facility codes at the point of data entry rather than relying on free-text names would remove a linkage hazard that produced 30 distinct spellings of nine facilities.

For research using these data, three implications follow. Analyses of mortality, prematurity severity and birth-weight classification can proceed with the caveats in table 5. Analyses of referral timing must be restricted to records with valid intervals and framed as birth-to-admission interval rather than transport delay. Analyses of documentation-derived quality scores must recognise that survivors have more opportunity to accumulate documented care than newborns who die, and should therefore use early-care rather than whole-admission exposures for mortality models. For counties, the broader point is that the fields that fail are not the fields most often audited, and a data-quality review examining only completeness of birth weight and outcome will report a system in better health than it is in.

## Conclusion

Routine maternity and newborn records in these Western Kenyan facilities were sufficiently complete and internally consistent to identify preterm and low-birth-weight newborns, classify prematurity severity, and analyse in-hospital mortality among newborns with a known final outcome. They were not sufficient to estimate preterm-birth prevalence, to characterise the referral and transport process, or to compute length of stay reliably from recorded dates. The dominant defects were chronological rather than biological, and the dominant gaps were in maternal history and timing fields rather than in the newborn’s clinical description. Mandatory timing fields with point-of-entry validation, automatic derivation of birth-weight categories, routine capture of facility-month delivery denominators, and coded facility identifiers would address most of what we found using systems that already exist. Because two of the nine facilities contributing newborn records accounted for a large share of the variation and the provider evidence comes from a single referral hospital, these findings describe the assessed facilities and should not be generalised to Western Kenya as a whole.

## Declarations

## Ethics approval and research authorisations

Ethical approval was granted by the Masinde Muliro University of Science and Technology Institutional Scientific and Ethics Review Committee (MMUST-ISERC), approval number MMUST/ISERC/023/2026 (correspondence reference MMU/COR: 40312 Vol 6(01)), dated 2 February 2026 and valid from 2 February 2026 to 2 February 2027, for the research proposal titled ‘Health Systems Readiness and Quality of Care for Preterm Births in Western Kenya’.

A national research licence was issued by the National Commission for Science, Technology and Innovation (NACOSTI), Kenya: licence number NACOSTI/P/26/4184987, reference number 168774, issued 17 February 2026 and valid until 17 February 2027, for research in Bungoma on the topic ‘Health Systems Readiness and Quality of Care for Preterm Births in Western Kenya’.

County research authorisation was granted by the County Government of Bungoma, Ministry of Health, Office of the County Director of Health, reference CG/BGM/CDH/RESRC/VOL.1, dated 24 February 2026.

## Consent to participate

MMUST-ISERC approved the retrospective abstraction of routine clinical records with the requested individual patient consent waiver. Providers who took part in key informant interviews gave informed consent to participate and to be audio-recorded, documented at the start of each interview. Identifiers were removed during abstraction, all reporting is aggregated, and quotations are anonymised by role.

## Patient consent for publication

No individual patient data, images or identifiable details are reported, so patient consent for publication was not required. Providers consented to the use of anonymised quotations from their interviews.

## Data availability statement

Data are available upon reasonable request. The de-identified analytical datasets, the derived-variable dictionary, the data-quality rule dictionary, the duplicate-resolution and record-lineage logs, and the analysis code are held by the study team at Kibabii University and may be made available to qualified researchers on reasonable request to the corresponding author, subject to approval by MMUST-ISERC, the County Government of Bungoma Department of Health and Kibabii University data-governance requirements, and to a data-sharing agreement. Individual patient-level clinical records will not be released publicly.

## Contributors

MSS: conceptualisation, methodology, formal analysis, data curation, writing – original draft. LNN: conceptualisation, supervision, investigation, writing – review and editing. CMS: conceptualisation, funding acquisition, project administration, investigation, writing – review and editing. TCK: methodology, validation, writing – review and editing. ENM: methodology, validation, writing – review and editing. MSS is the guarantor and accepts full responsibility for the work and the conduct of the study, had access to the data, and controlled the decision to publish. All named authors meet the ICMJE criteria for authorship, have approved the final manuscript and agree to be accountable for all aspects of the work.

## Data Availability

All data produced in the present study are available upon reasonable request to the authors, subject to approval by the Masinde Muliro University of Science and Technology Institutional Scientific and Ethics Review Committee and the County Government of Bungoma Department of Health. Individual patient-level clinical records will not be released publicly.

## Acknowledgements

The authors acknowledge the County Government of Bungoma Department of Health, the management and staff of the participating health facilities, the health records and information officers who supported record retrieval, the research assistants who conducted the abstraction, and the providers who gave their time to the key informant interviews. Lastly, the authors wish to acknowledge Pauline Lusaria Waswa who was the volunteer project administrator. Claude (Anthropic), a large language model, was used to assist with language editing and manuscript checks against journal requirements. The authors designed the study, collected and analysed the data, interpreted the findings, and reviewed, revised and take full responsibility for all content.

## Provenance and peer review

Not commissioned; externally peer reviewed.

## References

1. World Health Organization. Standards for improving the quality of care for small and sick newborns in health facilities. Geneva: World Health Organization, 2020.

2. Moxon SG, Ruysen H, Kerber KJ, et al. Count every newborn; a measurement improvement roadmap for coverage data. BMC Pregnancy Childbirth 2015;15(Suppl 2):S8.

3. Ohuma EO, Moller AB, Bradley E, et al. National, regional, and global estimates of preterm birth in 2020, with trends from 2010: a systematic analysis. Lancet 2023;402:1261–71.

4. Lawn JE, Ohuma EO, Bradley E, et al. Small babies, big risks: global estimates of prevalence and mortality for vulnerable newborns to accelerate change and improve counting. Lancet 2023;401:1707–19.

5. Kenya National Bureau of Statistics, Ministry of Health. Kenya Demographic and Health Survey 2022: main report, volume 1. Nairobi: KNBS, 2023.

6. Okwaraji YB, Krasevec J, Bradley E, et al. National routine data for low birthweight and preterm births: systematic data quality assessment for United Nations member states (2000–2020). BJOG 2024. doi:10.1111/1471-0528.17699.

7. Miller L, Wanduru P, Santos N, et al. Working with what you have: how the East Africa Preterm Birth Initiative used gestational age data from facility maternity registers. PLoS One 2020;15:e0237656.

8. Bucher S, Nowak K, Otieno K, et al. Birth weight and gestational age distributions in a rural Kenyan population. BMC Pediatr 2023;23:107.

9. Wagenaar BH, Sherr K, Fernandes Q, et al. Using routine health information systems for well-designed health evaluations in low-and middle-income countries. Health Policy Plan 2016;31:129–35.

10. Lundin R, Mariani I, Peven K, et al. Quality of routine health facility data used for newborn indicators in low-and middle-income countries: a systematic review. J Glob Health 2022;12:04019.

11. Kananura RM, Paolo D, Cora LG, et al. Availability and the quality of key newborn data within routine health facility data: findings of the IMPULSE observational study in the Central African Republic, Ethiopia, Tanzania, and Uganda. J Glob Health 2025;15:04359.

12. Odeny BM, Njoroge A, Gloyd S, et al. Development of novel composite data quality scores to evaluate facility-level data quality in electronic data in Kenya: a nationwide retrospective cohort study. BMC Health Serv Res 2023;23:1139.

13. Oware PM, Omondi G, Adipo C, et al. Perceived accuracy and utilisation of DHIS2 data for health decision making and advocacy in Kenya: a qualitative study. PLOS Glob Public Health 2025;5:e0004508.

14. Maïga A, Jiwani SS, Mutua MK, et al. Generating statistics from health facility data: the state of routine health information systems in Eastern and Southern Africa. BMJ Glob Health 2019;4:e001849.

15. Tuti T, Bitok M, Paton C, et al. Innovating to enhance clinical data management using non-commercial and open source solutions across a multi-center network supporting inpatient pediatric care and research in Kenya. J Am Med Inform Assoc 2016;23:184–92.

16. Irimu G, Aluvaala J, Malla L, et al. Neonatal mortality in Kenyan hospitals: a multisite, retrospective, cohort study. BMJ Glob Health 2021;6:e004475.

17. Ayieko P, Ogero M, Makone B, et al. Characteristics of admissions and variations in the use of basic investigations, treatments and outcomes in Kenyan hospitals within a new clinical information network. Arch Dis Child 2016;101:223–9.

18. Hagel C, Paton C, Mbevi G, et al. Data for tracking SDGs: challenges in capturing neonatal data from hospitals in Kenya. BMJ Glob Health 2020;5:e002108.

19. Kihuba E, Gathara D, Mwinga S, et al. Assessing the ability of health information systems in hospitals to support evidence-informed decisions in Kenya. Glob Health Action 2014;7:24859.

20. Chiba Y, Oguttu MA, Nakayama T. Quantitative and qualitative verification of data quality in the childbirth registers of two rural district hospitals in Western Kenya. Midwifery 2012;28:329–39.

21. Benchimol EI, Smeeth L, Guttmann A, et al. The REporting of studies Conducted using Observational Routinely-collected health Data (RECORD) statement. PLoS Med 2015;12:e1001885.

22. von Elm E, Altman DG, Egger M, et al. The Strengthening the Reporting of Observational Studies in Epidemiology (STROBE) statement: guidelines for reporting observational studies. Lancet 2007;370:1453–7.

23. Tong A, Sainsbury P, Craig J. Consolidated criteria for reporting qualitative research (COREQ): a 32-item checklist for interviews and focus groups. Int J Qual Health Care 2007;19:349–57.

24. World Health Organization. Data quality review: a toolkit for facility data quality assessment. Module 1: framework and metrics. Geneva: World Health Organization, 2017.

25. Aqil A, Lippeveld T, Hozumi D. PRISM framework: a paradigm shift for designing, strengthening and evaluating routine health information systems. Health Policy Plan 2009;24:217–28.

26. Bland JM, Altman DG. Statistical methods for assessing agreement between two methods of clinical measurement. Lancet 1986;1:307–10.

27. Zou G. A modified Poisson regression approach to prospective studies with binary data. Am J Epidemiol 2004;159:702–6.

28. Gale NK, Heath G, Cameron E, et al. Using the framework method for the analysis of qualitative data in multi-disciplinary health research. BMC Med Res Methodol 2013;13:117.

29. Newcombe RG. Two-sided confidence intervals for the single proportion: comparison of seven methods. Stat Med 1998;17:857–72.

30. Fleiss JL, Cohen J, Everitt BS. Large sample standard errors of kappa and weighted kappa. Psychol Bull 1969;72:323–7.

31. Keating R, Merai R, Mubiri P, et al. Assessing effects of a data quality strengthening campaign on completeness of key fields in facility-based maternity registers in Kenya and Uganda. East African Journal of Applied Health Monitoring and Evaluation 2019;3:20–8.

32. Shamba D, Day LT, Zaman SB, et al. Barriers and enablers to routine register data collection for newborns and mothers: EN-BIRTH multi-country validation study. BMC Pregnancy Childbirth 2021;21(Suppl 1):233.

33. Day LT, Sadeq-Ur Rahman Q, Ehsanur Rahman A, et al. Assessment of the validity of the measurement of newborn and maternal health-care coverage in hospitals (EN-BIRTH): an observational study. Lancet Glob Health 2021;9:e267–79.

34. Kong S, Day LT, Zaman SB, et al. Birthweight: EN-BIRTH multi-country study. BMC Pregnancy Childbirth 2021;21(Suppl 1):240.

35. Landry E, Pett C, Fiorentino R, et al. Assessing the quality of record keeping for cesarean deliveries: results from a multicenter retrospective record review in five low-income countries. BMC Pregnancy Childbirth 2014;14:139.

36. Day LT, Gore-Langton GR, Rahman AE, et al. Labour and delivery ward register data availability, quality, and utility — Every Newborn birth indicators research tracking in hospitals (EN-BIRTH) study baseline analysis in three countries. BMC Health Serv Res 2020;20:737.

37. Rumisha SF, Lyimo EP, Mremi IR, et al. Data quality of the routine health management information system at the primary healthcare facility and district levels in Tanzania. BMC Med Inform Decis Mak 2020;20:340.

38. Shama AT, Roba HS, Abaerei AA, et al. Assessment of quality of routine health information system data and associated factors among departments in public health facilities of Harari region, Ethiopia. BMC Med Inform Decis Mak 2021;21:287.

39. Kinney MV, Ajayi G, de Graft-Johnson J, et al. ’It might be a statistic to me, but every death matters’: an assessment of facility-level maternal and perinatal death surveillance and response systems in four sub-Saharan African countries. PLoS One 2020;15:e0243722.

40. Cetin K, Worku D, Demtse A, et al. ’Death audit is a fight’: provider perspectives on the ethics of the maternal and perinatal death surveillance and response (MPDSR) system in Ethiopia. BMC Health Serv Res 2022;22:1214.

41. Bhattacharya AA, Allen E, Umar N, et al. Improving the quality of routine maternal and newborn data captured in primary health facilities in Gombe State, Northeastern Nigeria: a before-and-after study. BMJ Open 2020;10:e038174.

42. Bhattacharya AA, Umar N, Audu A, et al. Quality of routine facility data for monitoring priority maternal and newborn indicators in DHIS2: a case study from Gombe State, Nigeria. PLoS One 2019;14:e0211265.

43. Muthee V, Bochner AF, Osterman A, et al. The impact of routine data quality assessments on electronic medical record data quality in Kenya. PLoS One 2018;13:e0195362.

44. Wainaina J, Ogero M, Mumelo L, et al. Hypothermia amongst neonatal admissions in Kenya: a retrospective cohort study assessing prevalence, trends, associated factors, and its relationship with all-cause neonatal mortality. Front Pediatr 2024;12:1272104.

