## Supplementary material for "Completeness, consistency and analytical usability of routine data for preterm and small newborn care in Western Kenya: a multicentre mixed-methods study": Online supplemental material

**Contents**

**Online supplemental table 1. Full data-quality rule dictionary (operational definition, fields involved and eligible denominator for all 41 prespecified rules)**

**Online supplemental table 2. Full variable-level missingness for every variable in both patient-level instruments**

**Online supplemental table 3. Facility-specific data-quality statistics**

**Online supplemental table 4. Duplicate-resolution results for both patient-level datasets**

**Online supplemental table 5. Sensitivity analyses**

**Online supplemental table 6. Joint display: integration of quantitative data-quality findings with facility context and provider accounts**

**Online supplemental figure 1. Maternity-register documentation completeness by variable and facility**

**Online supplemental figure 2. Bland–Altman analysis of duplicated gestational-age and birth-weight fields**

**Online supplemental figure 3. Mean record-level documentation completeness by facility and instrument**

**Online supplemental figure 4. Facility health-information-system and overall readiness against mean record-level documentation completeness**

**Online supplemental file 1. Completed RECORD checklist incorporating STROBE items for cross-sectional studies**

### Online supplemental table 1. Full data-quality rule dictionary

| Instrument | Dimension | Rule | Fields involved | Operational definition | Eligible n |
|---|---|---|---|---|---|
| Tool 1 (maternity register) | Internal consistency | Recorded preterm status agrees with gestational age (<37 weeks) | Preterm classification; gestational age (weeks) | Flagged where gestational age <37 with preterm recorded as No, or >=37 with preterm recorded as Yes | 340 |
| Tool 1 (maternity register) | Internal consistency | Recorded prematurity category agrees with gestational-age band | Prematurity category; gestational age | Flagged where the recorded category differs from the band derived from gestational age (<28, 28–31, 32–36, >=37 weeks) | 325 |
| Tool 1 (maternity register) | Internal consistency | “Birth weight not recorded” indicator agrees with entered birth weight | Birth-weight not-recorded switch; birth weight (g) | Flagged where the switch is Yes but a weight is entered, or No but no weight is entered | 262 |
| Tool 1 (maternity register) | Internal consistency | “Gravidity not recorded” indicator agrees with entered gravidity | Gravidity not-recorded switch; gravidity | Flagged where the switch and the entered value contradict each other | 266 |
| Tool 1 (maternity register) | Internal consistency | “Parity not recorded” indicator agrees with entered parity | Parity not-recorded switch; parity | Flagged where the switch and the entered value contradict each other | 265 |
| Tool 1 (maternity register) | Internal consistency | Parity does not exceed gravidity | Gravidity; parity | Flagged where parity > gravidity | 347 |
| Tool 1 (maternity register) | Internal consistency | “No maternal comorbidity recorded” agrees with individual comorbidity items | Comorbidity composite; individual comorbidity items | Flagged where None recorded is selected but at least one individual comorbidity is also selected | 297 |
| Tool 1 (maternity register) | Internal consistency | “Comorbidity not recorded” agrees with individual comorbidity items | Comorbidity not-recorded switch; individual comorbidity items | Flagged where Not recorded is selected but at least one individual comorbidity is also selected | 297 |
| Tool 1 (maternity register) | Internal consistency | Abstractor-reported key-variable completeness agrees with computed completeness | Abstractor key-variables-complete field; study record ID, delivery date, gestational age or preterm status, birth weight, newborn sex, birth outcome | Flagged where the abstractor answer differs from completeness computed from the six key fields | 349 |
| Tool 1 (maternity register) | Plausibility | Maternal age within 10–55 years | Maternal age (years) | Flagged outside 10–55 years | 346 |
| Tool 1 (maternity register) | Plausibility | Gestational age within 20–44 weeks | Gestational age (weeks) | Flagged outside 20–44 weeks | 341 |
| Tool 1 (maternity register) | Plausibility | Birth weight within 300–6000 g | Birth weight (g) | Flagged outside 300–6,000 g | 347 |
| Tool 1 (maternity register) | Plausibility | Gravidity within 1–20 | Gravidity | Flagged outside 1–20 | 348 |
| Tool 1 (maternity register) | Plausibility | Parity within 0–19 | Parity | Flagged outside 0–19 | 347 |
| Tool 1 (maternity register) | Plausibility | Delivery date not later than date of data collection | Delivery date; data-collection date | Flagged where delivery date is later than the date the record was abstracted (device submission date) | 349 |
| Tool 1 (maternity register) | Plausibility | Gestational age and birth weight biologically compatible | Gestational age; birth weight | Flagged where gestational age <28 weeks with weight >=2,000 g, or >=34 weeks with weight <800 g | 340 |
| Tool 3 (newborn record) | Internal consistency | Recorded preterm status agrees with gestational age (<37 weeks) | Preterm classification; gestational age | Flagged where gestational age <37 with preterm recorded as No, or >=37 with preterm recorded as Yes | 272 |
| Tool 3 (newborn record) | Internal consistency | Recorded low-birth-weight status agrees with birth weight (<2500 g) | LBW classification; birth weight | Flagged where weight <2,500 g with LBW recorded as No, or >=2,500 g with LBW recorded as Yes | 249 |
| Tool 3 (newborn record) | Internal consistency | Recorded very-low-birth-weight status agrees with birth weight (<1500 g) | VLBW classification; birth weight | Flagged where weight <1,500 g with VLBW recorded as No, or >=1,500 g with VLBW recorded as Yes | 228 |
| Tool 3 (newborn record) | Internal consistency | Duplicated gestational-age fields agree exactly | Two gestational-age fields | Flagged where the two recorded values differ | 238 |
| Tool 3 (newborn record) | Internal consistency | Duplicated birth-weight fields agree exactly | Two birth-weight fields | Flagged where the two recorded values differ | 259 |
| Tool 3 (newborn record) | Internal consistency | Admission weight not more than 10% above birth weight | Birth weight; admission weight | Flagged where admission weight exceeds 110% of birth weight | 242 |
| Tool 3 (newborn record) | Internal consistency | Recorded length of stay within 1 day of interval calculated from dates | Recorded length of stay; admission and outcome date-times | Flagged where the absolute difference exceeds one day | 225 |
| Tool 3 (newborn record) | Internal consistency | Admission date/time not earlier than birth date/time | Birth date-time; admission date-time | Flagged where admission precedes birth | 257 |

| Instrument | Dimension | Rule | Fields involved | Operational definition | Eligible n |
|---|---|---|---|---|---|
| Tool 3 (newborn record) | Internal consistency | Outcome date/time not earlier than admission date/time | Admission date-time; outcome date-time | Flagged where the outcome precedes admission | 236 |
| Tool 3 (newborn record) | Internal consistency | Disposition agrees with survival status at end of stay | Disposition; survival status | Flagged where Died is paired with Alive, or Discharged alive with Dead | 257 |
| Tool 3 (newborn record) | Internal consistency | “No complications recorded” agrees with individual complication items | Complications composite; individual complication items | Flagged where None recorded is selected alongside an individual complication | 270 |
| Tool 3 (newborn record) | Internal consistency | “No maternal infection” agrees with individual infection items | Maternal infection composite; individual infection items | Flagged where None is selected alongside an individual infection | 279 |
| Tool 3 (newborn record) | Internal consistency | Kangaroo mother care initiation scored only where eligibility assessment was applicable | KMC eligibility item; KMC initiation item | Flagged where eligibility is marked not applicable but initiation carries a numerical score | 291 |
| Tool 3 (newborn record) | Internal consistency | Feeding at discharge documented only for newborns discharged alive | Feeding at discharge; disposition | Flagged where feeding at discharge is completed for a newborn not discharged alive | 185 |
| Tool 3 (newborn record) | Internal consistency | Apgar “not recorded” indicator agrees with entered Apgar scores | Apgar not-recorded switch; 5-minute Apgar | Flagged where the switch and the entered score contradict each other | 250 |
| Tool 3 (newborn record) | Plausibility | Mother age within 10–55 years | Mother age (years) | Flagged outside 10–55 years | 226 |
| Tool 3 (newborn record) | Plausibility | Gestational age within 20–44 weeks | Gestational age (weeks) | Flagged outside 20–44 weeks | 275 |
| Tool 3 (newborn record) | Plausibility | Birth weight within 300–6000 g | Birth weight (g) | Flagged outside 300–6,000 g | 274 |
| Tool 3 (newborn record) | Plausibility | Apgar scores within 0–10 | 1-minute and 5-minute Apgar | Flagged outside 0–10 | 244 |
| Tool 3 (newborn record) | Plausibility | Birth-to-admission interval non-negative | Birth date-time; admission date-time | Flagged where the interval is negative | 257 |
| Tool 3 (newborn record) | Plausibility | Calculated length of stay non-negative | Admission and outcome date-times | Flagged where the calculated interval is negative | 236 |
| Tool 3 (newborn record) | Plausibility | Length of stay ≤180 days | Recorded length of stay | Flagged above 180 days | 247 |
| Tool 3 (newborn record) | Plausibility | Average daily weight change within −100 to +100 g/day | Admission weight; discharge weight; length of stay | Flagged outside −100 to +100 g per day | 209 |
| Tool 3 (newborn record) | Plausibility | Date of birth not later than date of data collection | Birth date; data-collection date | Flagged where the birth calendar date is later than the date the record was abstracted (device submission date) | 280 |
| Tool 3 (newborn record) | Plausibility | Outcome date not later than date of data collection | Outcome date; data-collection date | Flagged where the outcome calendar date is later than the date the record was abstracted (device submission date) | 259 |

*Rules are reproduced as prespecified and correspond one-to-one with the rows of Table 3 in the main text, which reports the result of each rule but not its operational definition. Eligible n is the number of records in which both elements of the rule were present, which is why denominators differ between rules.*

#### Online supplemental table 2. Full variable-level missingness

| Domain | Variable | Eligible n | Documented n | Blank n | Explicit 'not recorded' n | Complete % (95% CI) |
|---|---|---|---|---|---|---|
| Identification | Facility name | 349 | 349 | 0 | 0 | 100.0 (98.9 to 100.0) |
| Identification | Facility/register delivery number | 349 | 349 | 0 | 0 | 100.0 (98.9 to 100.0) |
| Identification | Date of delivery | 349 | 349 | 0 | 0 | 100.0 (98.9 to 100.0) |
| Maternal demographics | Maternal age (years) | 349 | 346 | 3 | 0 | 99.1 (97.5 to 99.7) |
| Maternal demographics | Residence type | 349 | 327 | 0 | 22 | 93.7 (90.6 to 95.8) |
| ANC and obstetric | ANC visits attended | 349 | 343 | 5 | 1 | 98.3 (96.3 to 99.2) |
| ANC and obstetric | ANC booking trimester | 349 | 298 | 16 | 35 | 85.4 (81.3 to 88.7) |
| ANC and obstetric | Gravidity | 349 | 348 | 1 | 0 | 99.7 (98.4 to 99.9) |
| ANC and obstetric | Parity | 349 | 347 | 2 | 0 | 99.4 (97.9 to 99.8) |
| ANC and obstetric | Multiple pregnancy | 349 | 343 | 6 | 0 | 98.3 (96.3 to 99.2) |
| ANC and obstetric | Mode of delivery | 349 | 348 | 1 | 0 | 99.7 (98.4 to 99.9) |
| ANC and obstetric | Maternal comorbidity status | 349 | 259 | 52 | 38 | 74.2 (69.4 to 78.5) |
| Newborn baseline | Gestational age (weeks) | 349 | 341 | 8 | 0 | 97.7 (95.5 to 98.8) |
| Newborn baseline | Gestational age source | 349 | 343 | 5 | 1 | 98.3 (96.3 to 99.2) |
| Newborn baseline | Birth weight (g) | 349 | 347 | 2 | 0 | 99.4 (97.9 to 99.8) |
| Newborn baseline | Sex of newborn | 349 | 345 | 3 | 1 | 98.9 (97.1 to 99.6) |
| Newborn baseline | Preterm classification | 349 | 348 | 0 | 1 | 99.7 (98.4 to 99.9) |
| Newborn baseline | Prematurity category | 341 | 325 | 16 | 0 | 95.3 (92.5 to 97.1) |
| Outcome | Birth outcome | 349 | 349 | 0 | 0 | 100.0 (98.9 to 100.0) |
| Identification | Facility name | 291 | 291 | 0 | 0 | 100.0 (98.7 to 100.0) |
| Identification | Newborn file number/MRN | 291 | 290 | 1 | 0 | 99.7 (98.1 to 99.9) |
| Identification | Date/time of birth | 291 | 280 | 11 | 0 | 96.2 (93.4 to 97.9) |
| Identification | Date/time of admission | 291 | 258 | 33 | 0 | 88.7 (84.5 to 91.8) |
| Maternal and obstetric | Mother age (years) | 291 | 226 | 65 | 0 | 77.7 (72.5 to 82.1) |
| Maternal and obstetric | Gravidity | 291 | 226 | 65 | 0 | 77.7 (72.5 to 82.1) |
| Maternal and obstetric | Parity | 291 | 221 | 70 | 0 | 75.9 (70.7 to 80.5) |
| Maternal and obstetric | ANC attendance | 291 | 204 | 13 | 74 | 70.1 (64.6 to 75.1) |
| Maternal and obstetric | ANC booking trimester | 291 | 163 | 35 | 93 | 56.0 (50.3 to 61.6) |
| Maternal and obstetric | Multiple pregnancy | 291 | 267 | 12 | 12 | 91.8 (88.0 to 94.4) |
| Maternal and obstetric | Mode of delivery | 291 | 281 | 10 | 0 | 96.6 (93.8 to 98.1) |
| Maternal and obstetric | Antenatal corticosteroids | 291 | 242 | 19 | 30 | 83.2 (78.4 to 87.0) |
| Newborn baseline | Gestational age (weeks) | 291 | 275 | 16 | 0 | 94.5 (91.3 to 96.6) |
| Newborn baseline | Gestational age source | 291 | 265 | 14 | 12 | 91.1 (87.2 to 93.8) |
| Newborn baseline | Birth weight (g) | 291 | 274 | 17 | 0 | 94.2 (90.8 to 96.3) |
| Newborn baseline | Preterm classification | 291 | 278 | 12 | 1 | 95.5 (92.5 to 97.4) |
| Newborn baseline | LBW classification | 291 | 254 | 35 | 2 | 87.3 (83.0 to 90.6) |
| Newborn baseline | VLBW classification | 291 | 233 | 58 | 0 | 80.1 (75.1 to 84.3) |
| Entry and referral | Place of birth | 291 | 291 | 0 | 0 | 100.0 (98.7 to 100.0) |

| Domain | Variable | Eligible n | Documented n | Blank n | Explicit ‘not recorded’ n | Complete % (95% CI) |
|---|---|---|---|---|---|---|
| Entry and referral | Mode of entry | 291 | 291 | 0 | 0 | 100.0 (98.7 to 100.0) |
| Entry and referral | Referring facility level | 60 | 58 | 2 | 0 | 96.7 (88.6 to 99.1) |
| Entry and referral | Distance/time to facility | 291 | 2 | 280 | 9 | 0.7 (0.2 to 2.5) |
| Admission assessment | Apgar at 1 minute | 291 | 244 | 47 | 0 | 83.8 (79.2 to 87.6) |
| Admission assessment | Apgar at 5 minutes | 291 | 244 | 47 | 0 | 83.8 (79.2 to 87.6) |
| Admission assessment | Hypothermia on admission | 291 | 266 | 15 | 10 | 91.4 (87.6 to 94.1) |
| Admission assessment | Hypoglycaemia on admission | 291 | 256 | 14 | 21 | 88.0 (83.7 to 91.2) |
| Admission assessment | Respiratory distress | 291 | 273 | 12 | 6 | 93.8 (90.4 to 96.1) |
| Admission assessment | Suspected sepsis | 291 | 273 | 11 | 7 | 93.8 (90.4 to 96.1) |
| Outcome | Disposition | 291 | 283 | 8 | 0 | 97.3 (94.7 to 98.6) |
| Outcome | Survival status at end of stay | 291 | 257 | 20 | 14 | 88.3 (84.1 to 91.5) |
| Outcome | Date/time of outcome | 291 | 259 | 32 | 0 | 89.0 (84.9 to 92.1) |
| Outcome | Length of stay (days) | 291 | 247 | 44 | 0 | 84.9 (80.3 to 88.5) |
| Discharge | Admission weight (g) | 291 | 248 | 43 | 0 | 85.2 (80.7 to 88.8) |
| Discharge | Discharge weight (g) | 174 | 169 | 5 | 0 | 97.1 (93.5 to 98.8) |
| Discharge | Feeding at discharge | 174 | 166 | 6 | 2 | 95.4 (91.2 to 97.7) |

*Maternity-register variables are listed first, followed by newborn-record variables. This table extends Table 2 in the main text by presenting blank and explicit ‘not recorded’ counts separately for every core variable in both instruments. Denominators exclude structurally inapplicable records.*

### Online supplemental table 3. Facility-specific data-quality statistics

| Facility | Level | T1 records | T1 mean completeness % | T1 complete core % | T1 inconsistency % | T1 duplicate % | T3 records | T3 mean completeness % | T3 complete core % | T3 inconsistency % | T3 duplicate % | HIS readines % |
|---|---|---|---|---|---|---|---|---|---|---|---|---|
| Bumula Sub-County Hospital | 4 | 15 | 99.3 | 86.7 | 13.3 | 0.0 | 3 | 99.0 | 66.7 | 66.7 | 0.0 | 62.5 |
| Bungoma County Referral Hospital | 5 | 77 | 99.0 | 84.4 | 13.0 | 0.0 | 82 | 93.4 | 26.8 | 75.6 | 2.4 | 100.0 |
| Cheptais Sub-County Hospital | | 23 | 96.6 | 39.1 | 13.0 | 0.0 | 23 | 93.7 | 8.7 | 26.1 | 0.0 | |
| Kabula Health Centre | 3 | 30 | 97.7 | 60.0 | 6.7 | 10.0 | 29 | 94.2 | 24.1 | 51.7 | 3.4 | 75.0 |
| Kimilili Sub-County Hospital | | 47 | 97.3 | 55.3 | 2.1 | 0.0 | 53 | 78.7 | 0.0 | 43.4 | 0.0 | |
| Mechimeru Model Health Centre | 3 | 18 | 93.1 | 27.8 | 38.9 | 0.0 | 1 | 93.9 | 0.0 | 100.0 | 0.0 | 50.0 |
| Naitiri Sub-County Hospital | 4 | 36 | 93.1 | 0.0 | 2.8 | 0.0 | 16 | 34.9 | 0.0 | 18.8 | 0.0 | 75.0 |
| Sirisia Sub-County Hospital | 4 | 17 | 98.8 | 82.4 | 70.6 | 5.9 | 2 | 100.0 | 100.0 | 0.0 | 0.0 | 87.5 |
| Webuye County Hospital | 4 | 86 | 95.3 | 29.1 | 20.9 | 7.0 | 82 | 96.2 | 51.2 | 46.3 | 1.2 | 100.0 |

*T1, maternity register; T3, newborn clinical record; HIS, health information system; QI, quality improvement. Readiness scores are blank for the two facilities that contributed patient-level records but no facility readiness assessment. Duplicate percentage is the proportion of that facility's source records excluded as confirmed duplicates.*

### Online supplemental table 4. Duplicate-resolution results

| Dataset | Group | Facility | Source record identifiers | Classification | Evidence used | Completeness scores | Record retained | Records excluded | Information recovered | Variables recovered | Conflicting variables | Resolution rule |
|---|---|---|---|---|---|---|---|---|---|---|---|---|
| T1 | T1D01 | Webuye County Hospital | T1-99-747849075;T1-72-747260318 | Confirmed duplicate | Matching facility, record identifiers, date and newborn characteristics | T1-99-747849075:100;T1-72-747260318:97.273 | T1-99-747849075 | T1-72-747260318 | Yes | other_comorb_text | abstraction_date;anc_booking_code;anc_booking_raw;caesarean;delivery_mode;delivery_within_period;early_anc_booking;issue_summary;mat_any_comorb;mat_comorb_count;mat_other_comorb;parity;period_to | Retain highes weighted-completeness record and recover only singular non-conflicting values |
| T1 | T1D02 | Webuye County Hospital | T1-97-747849061;T1-73-747260336 | Confirmed duplicate | Matching facility, record identifiers, date and newborn characteristics | T1-97-747849061:100;T1-73-747260336:100 | T1-97-747849061 | T1-73-747260336 | No | | abstraction_date;birth_weight_g;issue_summary | Retain highes weighted-completeness record and recover only singular non-conflicting values |
| T1 | T1D03 | Webuye County Hospital | T1-74-747264918;T1-95-747787597 | Confirmed duplicate | Matching facility, record identifiers, date and newborn characteristics | T1-74-747264918:100;T1-95-747787597:100 | T1-74-747264918 | T1-95-747787597 | Yes | other_comorb_text;register_notes | abstraction_date;anc_booking_code;anc_booking_raw;caesarean;delivery_mode;early_anc_booking;mat_any_comorb;mat_comorb_count;mat_other_comorb | Retain highes weighted-completeness record and recover only singular non-conflicting values |
| T1 | T1D04 | Webuye County Hospital | T1-75-747270751;T1-96-747789095 | Confirmed duplicate | Matching facility, record identifiers, date and newborn characteristics | T1-75-747270751:100;T1-96-747789095:100 | T1-75-747270751 | T1-96-747789095 | No | | abstraction_date;anc_4plus;anc_visits_code;anc_visits_raw;caesarean;delivery_mode;other_comorb_text;parity;residence_name | Retain highes weighted-completeness record and recover only singular non-conflicting values |
| T1 | T1D05 | Webuye County Hospital | T1-94-747785858;T1-76-747275160 | Confirmed duplicate | Matching facility, record identifiers, date and newborn characteristics | T1-94-747785858:100;T1-76-747275160:97.273 | T1-94-747785858 | T1-76-747275160 | Yes | register_notes | abstraction_date;anc_4plus;anc_visits_code;anc_visits_raw;gravidity;maternal_age;nulliparous;parity;primigravida | Retain highes weighted-completeness record and recover only singular non-conflicting values |
| T1 | T1D06 | Webuye County Hospital | T1-89-747740603;T1-92-747782303 | Confirmed duplicate | Matching facility, record identifiers, date and newborn characteristics | T1-89-747740603:100;T1-92-747782303:100 | T1-89-747740603 | T1-92-747782303 | No | | gravidity;other_comorb_text;parity;residence_name | Retain highes weighted-completeness record and recover only singular non-conflicting values |
| T1 | T1D07 | Sirisia Sub-County Hospital | T1-228-750940109;T1-229-750945387 | Confirmed duplicate | Matching facility, record identifiers, date and newborn characteristics | T1-228-750940109:100;T1-229-750945387:96.071 | T1-228-750940109 | T1-229-750945387 | No | | core_complete;core_missing_count;facility_name_raw;issue_summary;newborn_sex | Retain highes weighted-completeness record and recover only singular non-conflicting values |

| Dataset | Group | Facility | Source record identifiers | Classification | Evidence used | Completeness scores | Record retained | Records excluded | Information recovered | Variables recovered | Conflicting variables | Resolution rule |
|---|---|---|---|---|---|---|---|---|---|---|---|---|
| T1 | T1D08 | Kabula Health Centre | T1-322-784580071;T1-347-793385765 | Confirmed duplicate | Matching facility, record identifiers, date and newborn characteristics | T1-322-784580071:100;T1-347-793385765:100 | T1-322-784580071 | T1-347-793385765 | No | | abstraction_date;analysis_id;mat_any_comorb;mat_any_infection; mat_comorb_count;mat_not_recorded;mat_uti_inf;nulliparous;parity;period_from;period_to;study_record_id_raw;subcounty_raw | Retain highes weighted-completeness record and recover only singular non-conflicting values |
| T1 | T1D09 | Kabula Health Centre | T1-323-784599519;T1-345-793311137 | Confirmed duplicate | Matching facility, record identifiers, date and newborn characteristics | T1-323-784599519:100;T1-345-793311137:96.071 | T1-323-784599519 | T1-345-793311137 | No | | abstraction_date;analysis_id;facility_name_raw;mat_any_comorb; mat_any_infection;mat_comorb_count;mat_none_recorded;mat_uti_inf;period_from;period_to;residence_name;study_record_id_raw;subcounty_raw | Retain highes weighted-completeness record and recover only singular non-conflicting values |
| T1 | T1D10 | Kabula Health Centre | T1-337-789385318;T1-338-789391426 | True multiple birth | Different newborn characteristics and explicit multiple-birth documentation | T1-337-789385318:100;T1-338-789391426:100 | All | | No | | Expected newborn-specific differences | Retain all tru multiple birth |
| T1 | T1D11 | Kabula Health Centre | T1-339-789460677;T1-342-793249997 | Confirmed duplicate | Matching facility, record identifiers, date and newborn characteristics | T1-339-789460677:100;T1-342-793249997:100 | T1-339-789460677 | T1-342-793249997 | No | | abstraction_date;analysis_id;facility_name_raw;residence_name;study_record_id_raw | Retain highes weighted-completeness record and recover only singular non-conflicting values |

| Dataset | Group | Facility | Source record identifiers | Classification | Evidence used | Completeness scores | Record retained | Records excluded | Information recovered | Variables recovered | Conflicting variables | Resolution rule |
|---|---|---|---|---|---|---|---|---|---|---|---|---|
| T3 | T3D01 | Webuye County Hospital | 161611-746737350;161611-746716835 | Confirmed duplicate | Matching facility, record identifiers, date and newborn characteristics | 161611-746737350:89.87;161611-746716835:85.325 | 161611-746737350 | 161611-746716835 | Yes | admission_datetime;admission_datetime_raw;birth_to_admission_hours;corrected_ga_discharge_calc;los_category;los_final_days;los_from_dates;major_doc_gaps;outcome_datetime;outcome_datetime_derived;outcome_datetime_raw;referral_delay_cat | admission_datetime;admission_datetime_raw;admission_instability_count;analysis_uid;birth_to_admission_hours;continuing_plqs_max;continuing_plqs_pct;continuing_plqs_score;corrected_ga_discharge_calc;early_plqs_max;early_plqs_pct;early_plqs_score;exposure_items_observed;inf_none;inf_unknown;issue_summary;los_final_days;los_from_dates;outcome_datetime;outcome_datetime_derived;outcome_datetime_raw;plqs_pct;plqs_quartile;plqs_total_max;plqs_total_score;q3_spo2;q3_vitals;q4_antibiotic_dose;q4_antibiotic_ind;q5_feeding_plan;q6_kmc_counselling;q6_kmc_duration;q8_escalation;q8_referral_stabilize;q_d3_pct;q_d4_max;q_d4_score;q_d5_pct;q_d5_score;q_d6_pct;q_d6_score;q_d8_max;q_d8_pct;q_d8_score;quality_full_items;referral_delay_cat | Retain highes weighted-completeness record and recover only singular non-conflicting values |

| Dataset | Group | Facility | Source record identifiers | Classification | Evidence used | Completeness scores | Record retained | Records excluded | Information recovered | Variables recovered | Conflicting variables | Resolution rule |
|---|---|---|---|---|---|---|---|---|---|---|---|---|
| T3 | T3D02 | Bungoma County Referral Hospital | 83-771555030;53-758168823 | Confirmed duplicate | Matching facility, record identifiers, date and newborn characteristics | 83-771555030:91.818;53-758168823:89.545 | 83-771555030 | 53-758168823 | Yes | corrected_ga_discharge_calc;los_category;los_final_days;los_from_dates;outcome_datetime;outcome_datetime_derived;outcome_datetime_raw | abstraction_date;abstractor_name;adm_hypoglycemia;admission_datetime;admission_datetime_raw;admission_instability_count;analysis_uid;avg_daily_weight_gain_g;birth_to_admission_hours;comp_jaundice;comp_rds;continuing_plqs_max;continuing_plqs_pct;continuing_plqs_score;corrected_ga_discharge_calc;discharge_weight_g;early_plqs_pct;early_plqs_score;facility_name_raw;ga_source;inf_none;inf_unknown;issue_summary;los_final_days;los_from_dates;major_doc_gaps;mother_age;multiple_pregnancy;outcome_datetime;outcome_datetime_derived;outcome_datetime_raw;plqs_pct;plqs_total_max;plqs_total_score;q2_clinical_review;q3_oxygen;q3_resuscitation;q3_spo2;q4_antibiotic_ind;q4_iv_fluids;q6_kmc_24h;q6_kmc_counselling;q6_kmc_duration;q7_hand_asepsis;q7_infection_monitor;q7_line_care;q8_escalation;q8_referral_stabilize;q_d2_pct;q_d2_score;q_d6_pct;q_d6_score;q_d7_pct;q_d7_score;q_d8_max;q_d8_pct;q_d8_score;quality_full_items;quality_zero_items;record_source_count;src_other;study_id;weight_change_final_g;weight_change_from_adm_g;weight_change_from_birth_g;weight_gain_positive | Retain highes weighted-completeness record and recover only singular non-conflicting values |

| Dataset | Group | Facility | Source record identifiers | Classification | Evidence used | Completeness scores | Record retained | Records excluded | Information recovered | Variables recovered | Conflicting variables | Resolution rule |
|---|---|---|---|---|---|---|---|---|---|---|---|---|
| T3 | T3D03 | Bungoma County Referral Hospital | 77-771410833;73-767237673 | Confirmed duplicate | Matching facility, record identifiers, date and newborn characteristics | 77-771410833:91.818;73-767237673:88.409 | 77-771410833 | 73-767237673 | No | | abstraction_date;admission_datetime;admission_datetime_raw;admission_weight_g;analysis_uid;avg_daily_weight_gain_g;baseline_weight_g;birth_to_admission_hours;birth_weight_disagreement;continuing_plqs_max;continuing_plqs_pct;continuing_plqs_score;corrected_ga_discharge_calc;early_plqs_max;early_plqs_pct;early_plqs_score;feeding_at_discharge;ga_source;issue_summary;los_final_days;los_from_dates;los_recorded_days;major_doc_gaps;outcome_datetime;outcome_datetime_raw;plqs_ge75;plqs_pct;plqs_quartile;plqs_total_max;plqs_total_score;q3_admission_assess;q3_oxygen;q3_spo2;q4_antibiotic_ind;q5_feeding_timely;q5_tolerance;q7_infection_monitor;q8_discharge_summary;q8_escalation;q8_referral_stabilize;q_d3_max;q_d3_pct;q_d3_score;q_d4_pct;q_d4_score;q_d5_pct;q_d5_score;q_d7_pct;q_d7_score;q_d8_max;q_d8_pct;q_d8_score;quality_full_items;src_other;src_referral;study_id;weight_change_final_g;weight_change_from_adm_g;weight_gain_positive | Retain highes weighted-completeness record and recover only singular non-conflicting values |

| Dataset | Group | Facility | Source record identifiers | Classification | Evidence used | Completeness scores | Record retained | Records excluded | Information recovered | Variables recovered | Conflicting variables | Resolution rule |
|---|---|---|---|---|---|---|---|---|---|---|---|---|
| T3 | T3D04 | Kabula Health Centre | 1591219-793245895;1591217-789448176 | Confirmed duplicate | Matching facility, record identifiers, date and newborn characteristics | 1591219-793245895:96.364;1591217-789448176:96.364 | 1591219-793245895 | 1591217-789448176 | Yes | exp_ipv;exposure_any;occupation | abstraction_date;admission_datetime;admission_datetime_raw;analysis_uid;avg_daily_weight_gain_g;birth_to_admission_hours;continuing_plqs_pct;continuing_plqs_score;corrected_ga_discharge_calc;discharge_weight_g;early_plqs_max;early_plqs_pct;early_plqs_score;education;exposure_items_observed;facility_name_raw;gravidity;inf_none;inf_unknown;issue_summary;los_category;los_final_days;los_from_dates;los_recorded_days;outcome_datetime;outcome_datetime_derived;outcome_datetime_raw;parity;plqs_pct;plqs_quartile;plqs_total_max;plqs_total_score;primigravida;q1_incubator_temp;q3_resuscitation;q4_iv_fluids;q4_vitamin_k;q8_escalation;q8_referral_stabilize;q_d1_pct;q_d1_score;q_d3_max;q_d3_pct;q_d4_max;q_d4_pct;q_d4_score;q_d8_pct;q_d8_score;quality_full_items;quality_zero_items;record_source_count;src_discharge;study_id;weight_change_final_g;weight_change_from_adm_g;weight_change_from_birth_g | Retain highes[illegible] weighted-completeness record and recover only singular non-conflicting values |

*Every duplicate group identified in both patient-level datasets, with the evidence used, the record retained, records excluded, variables recovered from complementary records, variables in conflict and the resolution rule applied. All exclusions are reversible through the record-lineage file.*

#### Online supplemental table 5. Sensitivity analyses

| Analysis | Dataset | Measure | Value |
|---|---|---|---|
| Primary (all source records) | Tool 1 | Mean record completeness % | 96.7 |
| Primary (all source records) | Tool 1 | Complete core record % | 50.1 |
| Restricted to unique records after duplicate resolution | Tool 1 | Mean record completeness % | 96.7 |
| Restricted to unique records after duplicate resolution | Tool 1 | Complete core record % | 50.3 |
| Primary (all source records) | Tool 3 | Mean record completeness % | 88.5 |
| Primary (all source records) | Tool 3 | Complete core record % | 26.5 |
| Restricted to unique records after duplicate resolution | Tool 3 | Mean record completeness % | 88.3 |
| Restricted to unique records after duplicate resolution | Tool 3 | Complete core record % | 25.9 |
| Any entry present counted as documented (explicit "not recorded"/"unknown" counted as documented) | Tool 1 | Mean record completeness % | 99.1 |
| Any entry present counted as documented (explicit "not recorded"/"unknown" counted as documented) | Tool 3 | Mean record completeness % | 90.3 |
| Restricted to facilities contributing ≥10 records | Tool 1 | Mean record completeness % | 96.7 |
| Restricted to facilities contributing ≥10 records | Tool 3 | Mean record completeness % | 88.3 |
| Restricted to facilities contributing ≥10 records | Tool 1 | Complete core record % | 50.1 |
| Restricted to facilities contributing ≥10 records | Tool 3 | Complete core record % | 25.6 |
| Excluding newborn records with chronologically invalid date sequences | Tool 3 | Mean record completeness % | 87.7 |
| Excluding newborn records with chronologically invalid date sequences | Tool 3 | Complete core record % | 27.6 |

*Primary analyses use all source records and count only usable entries as documented. The 'any entry' analysis counts explicit 'not recorded' and 'unknown' responses as documented, and is presented to separate genuine blank space from explicit negative responses.*

#### Online supplemental table 6. Joint display: integration of quantitative data-quality findings with facility context and provider accounts

| Quantitative data-quality finding | Facility HIS/QI context | Provider explanation | Clinical or programmatic implication | Recommended corrective action | Integration |
|---|---|---|---|---|---|
| Maternal history documented in 56–78% of newborn records; entire maternal domain blank in all 53 records at one hospital | HIS readiness 50–100%; six of ten facilities scored 12.5% on QI and governance | 'We normally miss each other. We normally focus on, is this baby sick? Yes. Is the baby out of danger? Yes.' (Provider KII 2) | Maternal and obstetric risk factors cannot be modelled for a quarter of newborns; confounding control is weakened | Link the maternity record to the newborn record at admission using a shared mother–newborn identifier rather than re-abstracting maternal history | Convergence |
| Recorded and date-derived length of stay differ by more than one day in 40.9% of records; admission precedes birth in 13.6% | Monthly data review conducted at the referral hospital via the Clinical Information Network | 'There is data review, which is done monthly… when that data is having errors, I'll be called to clarify.' (Provider KII 2) | Referral interval and length-of-stay indicators cannot be computed reliably from recorded dates | Add point-of-entry date-sequence validation so that an admission preceding birth cannot be saved; add date checks to the monthly review | Discordance |
| Vital-sign and monitoring items scored partial or absent in a substantial minority of records | Staff shortage named as a major gap by nine of ten facilities | 'We also monitor vitals hourly. That is the bad thing. Hourly with the shortage, sometimes we miss, sometimes we do it.' (Provider KII 2) | Documented monitoring frequency reflects staffing as much as clinical need, so quality scores partly measure workload | Interpret monitoring-derived quality items alongside nurse-to-newborn ratios; do not treat them as pure care indicators | Convergence |
| Records for newborns who died were less complete (adjusted PR 0.46, 95% CI 0.25 to 0.84) | Perinatal death review held on a fixed monthly cycle at the referral hospital | 'Every third Friday of the month is MPDSR. So it's permanent.' (Provider KII 1) | Documentation-derived quality exposures are subject to survivor time bias for mortality analysis | Use early-care rather than whole-admission quality exposures in mortality models; complete a minimum dataset at the time of death review | Convergence |
| Two birth-weight fields disagree in 10.0% of records, with differences up to 1,725 g; two gestational-age fields disagree in 8.8% | Structured paper forms transcribed to REDCap after discharge | 'As we take care of the baby, after discharging the baby, they take the patient file and enter that information.' (Provider KII 1) | Repeated capture detects error but does not resolve it; a small minority of newborns could be misclassified by weight category | Auto-populate the second field from the first, or flag discrepancies for resolution before submission | Expansion |
| Thirty distinct facility-name strings resolved to nine facilities; 15 sub-county spellings to seven sub-counties | All facilities report to KHIS; KHIS facility codes exist for all Bungoma facilities | Not raised by providers | Cross-dataset linkage and county aggregation are at risk of silent mis-assignment | Replace free-text facility entry with a coded KHIS facility list in the data-collection instrument | Expansion |
| Recorded LBW flag under-identified 15 of 238 newborns weighing below 2,500 g (exact McNemar p=0.001) | Gestational-age and birth-weight documentation scored 2 in nine of ten facilities | Not raised by providers | Low-birth-weight newborns may be missed for KMC eligibility, follow-up and reporting | Derive LBW and VLBW automatically from recorded weight instead of capturing them as separate items | Expansion |
| Register documentation is strong (median 100%) while newborn-record documentation is weaker (median 93.5%) and far more variable | Referral hospital has the highest readiness (83.1%) and highest patient volume | 'Luckily, our hospital is part of a network… This network has computers installed.' (Provider KII 1) | Investment in register and reporting systems does not automatically improve individual clinical documentation | Extend structured forms and validation from the register to the bedside clinical record | Complementarity |
| Audit recommendations recur without resolution across cycles | QI team and review meetings absent in six of ten facilities | 'Our only weakness has been in implementation of actual plans because some gaps keep recurring.' (Provider KII 1) | Audit generates findings without measurable change, and data-quality defects persist across review cycles | Assign a named owner and review date to each audit action; report closure rate as a standing QI indicator | Complementarity |

*HIS, health information system; QI, quality improvement; KII, key informant interview; PR, prevalence ratio; MPDSR, maternal and perinatal death surveillance and response; KMC, kangaroo mother care; LBW, low birth weight; VLBW, very low birth weight. Quotations are verbatim from the two provider interviews conducted at the county referral hospital.*

#### Online supplemental figure 1. Maternity-register documentation completeness by variable and facility

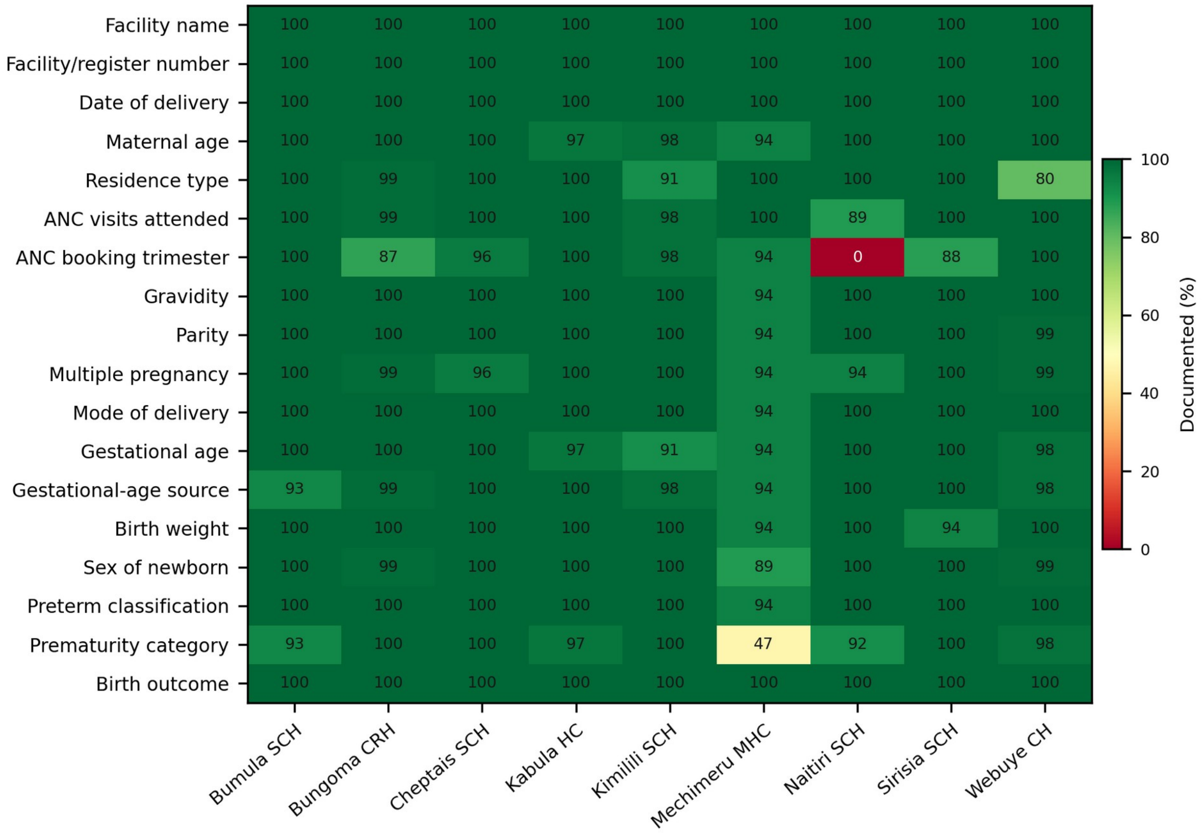


*Online supplemental figure 1. Documentation completeness of maternity-register records by variable and facility (Tool 1; 349 source records). Cell values are the percentage of that facility's records in which the variable carried a usable entry, computed from original uncleaned abstractions. The prematurity-category row is restricted to records with a recorded gestational age. CRH, county referral hospital; CH, county hospital; SCH, sub-county hospital; HC, health centre; MHC, model health centre.*

#### Online supplemental figure 2. Bland–Altman analysis of duplicated measurement fields

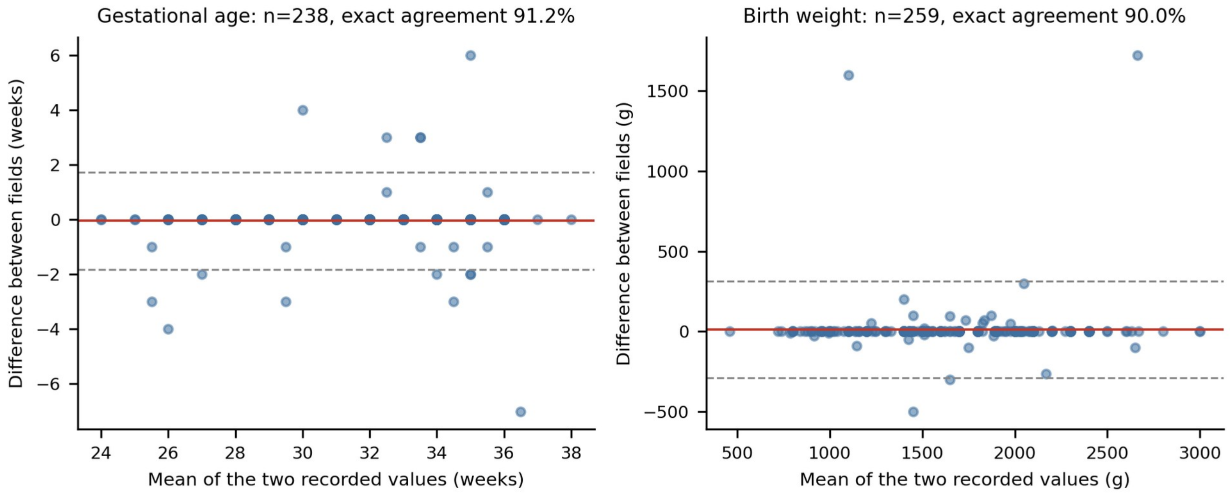


*Online supplemental figure 2. Bland–Altman analysis of the two fields capturing gestational age and the two fields capturing birth weight in the newborn clinical record. The solid line is the mean difference and the dashed lines the 95% limits of agreement. Exact agreement was 91.2% for gestational age and 90.0% for birth weight; the maximum absolute differences were 7 weeks and 1,725 g respectively.*

#### Online supplemental figure 3. Mean record-level documentation completeness by facility and instrument

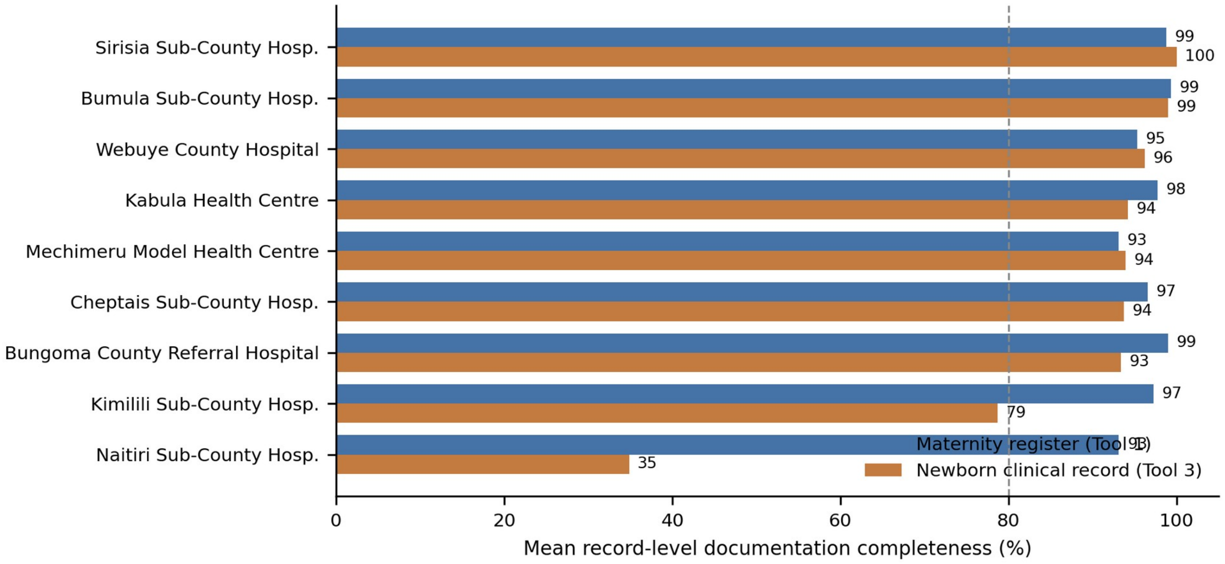


*Online supplemental figure 3. Mean record-level documentation completeness by facility and instrument. Bars show the mean proportion of applicable core fields documented per record for the maternity register (19 core fields) and the newborn clinical record (34 core fields, excluding distance or travel time). The dashed line marks 80%. Facilities are ordered by newborn-record completeness.*

#### Online supplemental figure 4. Facility readiness against mean record-level documentation completeness

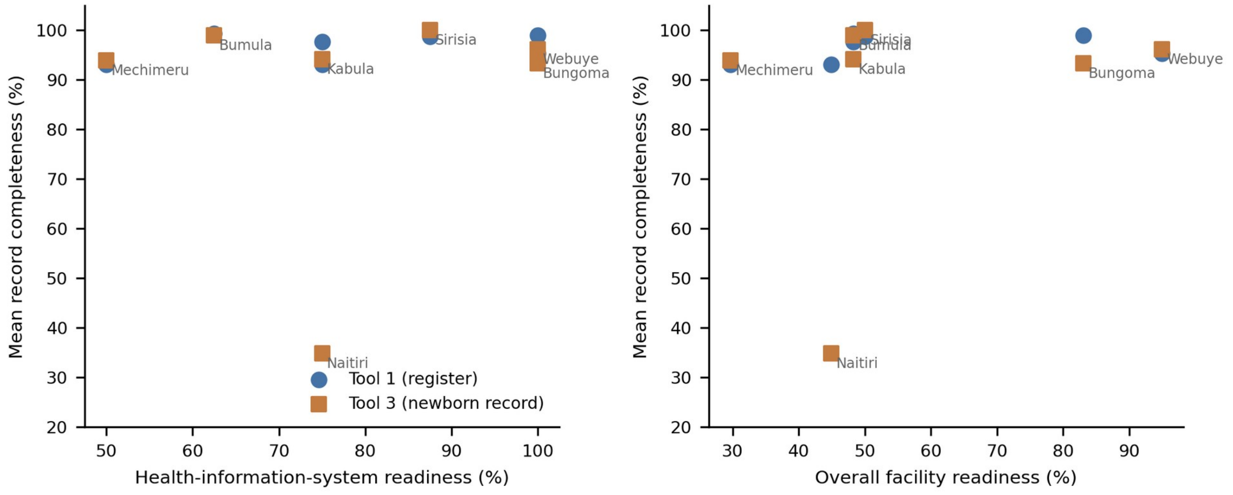


*Online supplemental figure 4. Facility health-information-system readiness and overall readiness against mean record-level documentation completeness. Each point is one of the seven facilities contributing both a readiness assessment and patient-level records. Spearman correlations were 0.27 (95% CI −0.64 to 0.87) for HIS readiness with register completeness and 0.04 (95% CI −0.76 to 0.79) for HIS readiness with newborn-record completeness. Relationships are ecological and exploratory.*

#### Online supplemental file 1. Completed RECORD checklist incorporating STROBE items

| Item | Requirement (abbreviated) | Location |
|---|---|---|
| STROBE 1a / RECORD 1.1 | Study design and databases indicated in title or abstract | Title; Abstract (Design; Participants) |
| RECORD 1.2 | Geographic region and timeframe stated | Title; Abstract (Setting; Participants); Results, Study records and facilities |
| RECORD 1.3 | Linkage between databases reported | Methods, Facility standardisation and duplicate resolution; Methods, Facility, HIS and predictor analyses |
| STROBE 1b | Structured abstract | Abstract |
| STROBE 2 | Scientific background and rationale | Introduction, paragraphs 1–5 |
| STROBE 3 | Objectives with prespecified hypotheses | Introduction, final paragraph |
| STROBE 4 | Study design presented early | Methods, Study design and reporting |
| STROBE 5 | Setting, locations and relevant dates | Methods, Setting; Methods, Data sources; Results, Study records and facilities |
| STROBE 6a / RECORD 6.1 | Eligibility, sources and selection; codes and algorithms | Methods, Data sources; Methods, Completeness assessment; online supplemental table 1 |
| RECORD 6.2 | Validation of codes or algorithms | Methods, Consistency, plausibility and agreement analysis (rules-based reference only; no external gold standard) |
| RECORD 6.3 | Person-level, institutional or other data linkage described | Methods, Facility standardisation and duplicate resolution; figure 1 |
| STROBE 7 / RECORD 7.1 | Variables, data sources and completeness of the databases | Methods, Data sources; Methods, Data-quality definitions; table 2 |
| STROBE 8 | Sources of data and measurement methods | Methods, Data sources; Methods, Data management |
| STROBE 9 | Efforts to address sources of bias | Methods, Data management; Methods, Missing data |
| STROBE 10 | Study size | Results, Study records and facilities; figure 1 |
| STROBE 11 | Quantitative variables and groupings | Methods, Completeness assessment; Methods, Consistency, plausibility and agreement analysis |
| STROBE 12a–e | Statistical methods, subgroups, missing data, sensitivity analyses | Methods, Facility, HIS and predictor analyses; Methods, Missing data; Methods, Statistical methods; online supplemental table 5 |
| RECORD 12.1 | Data cleaning methods described | Methods, Data management; Methods, Facility standardisation and duplicate resolution |
| RECORD 12.2 | Linkage quality evaluation | Results, Data verification and uniqueness |
| STROBE 13 / RECORD 13.1 | Participant flow diagram | Figure 1 |
| STROBE 14 | Descriptive data | Table 1; table 2 |
| STROBE 15 | Outcome data | Tables 2–4; figures 2–5 |

| Item | Requirement (abbreviated) | Location |
|---|---|---|
| STROBE 16 | Main results with precision | Tables 2–4 and 6; Results throughout |
| STROBE 17 | Other analyses | Results, Factors associated with complete documentation; online supplemental table 5 |
| STROBE 18 | Key results summarised | Discussion, Principal findings |
| STROBE 19 / RECORD 19.1 | Limitations, including those specific to routinely collected data | Discussion, Limitations |
| STROBE 20 | Interpretation | Discussion, Interpreting the completeness findings onwards |
| STROBE 21 | Generalisability | Discussion, Limitations; Conclusion |
| STROBE 22 / RECORD 22.1 | Funding; and access to the protocol, raw data and programming code | Funding; Data availability statement |
| COREQ (selected) | Research team, participant selection, setting, data collection, analysis and reporting for the embedded qualitative component | Methods, Data sources; Methods, Qualitative analysis; Results, Provider perspectives |

*RECORD, REporting of studies Conducted using Observational Routinely-collected health Data; STROBE, Strengthening the Reporting of Observational Studies in Epidemiology; COREQ, Consolidated Criteria for Reporting Qualitative Research. COREQ principles were applied selectively to the embedded explanatory qualitative component.*
